# Large Language Models and Medical Knowledge Grounding for Diagnosis Prediction

**DOI:** 10.1101/2023.11.24.23298641

**Authors:** Yanjun Gao, Ruizhe Li, Emma Croxford, Samuel Tesch, Daniel To, John Caskey, Brian W. Patterson, Matthew M. Churpek, Timothy Miller, Dmitriy Dligach, Majid Afshar

**Affiliations:** Department of Medicine, School of Medicine and Public Health, University of Wisconsin Madison; Department of Computing Science, University of Aberdeen; Department of Computer Science, Loyola University Chicago; Boston Children’s Hospital and Harvard Medical School

## Abstract

While Large Language Models (LLMs) have showcased their potential in diverse language tasks, their application in the healthcare arena needs to ensure the minimization of diagnostic errors and the prevention of patient harm. A Medical Knowledge Graph (KG) houses a wealth of structured medical concept relations sourced from authoritative references, such as UMLS, making it a valuable resource to ground LLMs’ diagnostic process in knowledge. In this paper, we examine the synergistic potential of LLMs and medical KG in predicting diagnoses given electronic health records (EHR), under the framework of Retrieval-augmented generation (RAG). We proposed a novel graph model: Dr.Knows, that selects the most relevant pathology knowledge paths based on the medical problem descriptions. In order to evaluate Dr.Knows, we developed the first comprehensive human evaluation approach to assess the performance of LLMs for diagnosis prediction and examine the rationale behind their decision-making processes, aimed at improving diagnostic safety. Using real-world hospital datasets, our study serves to enrich the discourse on the role of medical KGs in grounding medical knowledge into LLMs, revealing both challenges and opportunities in harnessing external knowledge for explainable diagnostic pathway and the realization of AI-augmented diagnostic decision support systems.

## 1 Introduction

The ubiquitous use of Electronic Health Records (EHRs) and the standard documentation practice of daily care notes are integral to the continuity of patient care by providing a comprehensive account of the patient’s health trajectory, inclusive of condition status, diagnoses, and treatment plans (Brown et al., 2014). Yet, the ever-increasing complexity and verbosity of EHR narratives, often laden with redundant information, presents the risk of cognitive overload for healthcare providers, potentially culminating in diagnostic inaccuracies (Rule et al., 2021; Liu et al., 2022; Nijor et al., 2022; Furlow, 2020) diagnostic inaccuracies (. Physicians often skip sections of lengthy and repetitive notes and rely on decisional shortcuts (i.e. decisional heuristics) that contribute to diagnostic errors (Croskerry, 2005).

Current efforts at automating diagnosis generation from daily progress notes leverage language models. Gao et al. (2022) introduced a summarization task that takes progress notes as input and generates a summary of active diagnoses. They annotated a set of progress notes from the publicly available EHR dataset called Medical Information Mart for Intensive Care III (mimic-iii) (Johnson et al., 2016). The BioNLP 2023 Shared Task, known as ProbSum, built upon this work by providing additional annotated notes and attracted multiple efforts focused on developing solutions Gao et al., 2023; Manakul et al., 2023; Li et al., 2023) These prior studies utilize language models like T5 (Raffel et al., 2020;) and GPT (Floridi and Chiriatti,2020), demonstrating a growing interest in applying generative large language models (LLMs) to serve as solutions. Unlike the conventional language tasks where LLMs have shown promising abilities, automated diagnosis generation is a critical task that requires high accuracy and reliability to ensure patient safety and optimize healthcare outcomes. Concerns regarding the potential misleading and hallucinated information that could result in life-threatening events prevent them from being utilized for diagnosis prediction (Baumgartner, 2023).

One of the solutions to improve factual accuracy is to utilize a knowledge graph to retrieve relevant knowledge to guide the LLMs with better instruction (Pan et al., 2023). In the biomedical domain, the Unified Medical Language System (UMLS) (Bodenreider, 2004), a comprehensive resource developed by the National Library of Medicine in the United States, has been extensively used in NLP research. It serves as the leading medical knowledge source, facilitating the integration and retrieval of biomedical information. The UMLS offers concept vocabulary and semantic relationships, enabling the construction of medical knowledge graphs. Prior studies have leveraged UMLS knowledge graphs for tasks such as information extraction (Huang et al., 2020; Lu et al., 2021; Aracena et al., 2022; He et al., 2020) and question-answering (Lu et al., 2021). However, UMLS knowledge graphs have not been applied to the task of diagnosis prediction.

Mining relevant knowledge for diagnosis is particularly challenging for two reasons: the highly specific factors related to the patient’s complaints, histories, and symptoms in EHR, and the vast search space within a knowledge graph containing 4.5 million concepts and 15 million relations for diagnosis determination. While utilizing a multi-hop reasoning mechanism for disease pathology discovery via the UMLS knowledge graph aligns with the need for extensive medical knowledge in diagnostics, implementing this approach is hampered by its computational complexity. Specifically, the number of concepts in the UMLS knowledge graph reachable within one hop ranges from 2 to 33k, with a median of 368. The number of two-hop paths may exhibit exponential growth due to the UMLS knowledge graph’s high connectivity. Therefore, addressing the computational complexity of multi-hop reasoning within the extensive UMLS knowledge graphs is crucial for effective knowledge mining in medical diagnostics.

In this study, we explore using knowledge graphs as an external module to ground LLM’s diagnostic process in medical knowledge and take the initial step of building a graph model to discover relevant paths using the UMLS. We propose Dr.Knows (**D**iagnostic **R**easoning **Know**ledge Graph**s**), that retrieves top N case-specific knowledge paths about the pathology of diseases through a multi-hop mechanism, overcoming the difficulties of retrieving and selecting paths from the entire knowledge graph. We then adapt the predicted paths into a graph-prompting method for LLMs. We utilized ChatGPT-3.5-turbo for our experiments on knowledge grounding since it represents the cutting-edge in language models and has been frequently examined as a diagnostic instrument in earlier research (Kuroiwa et al., 2023; Caruccio et al., 2024).

Going beyond the technical aspects of constructing knowledge graphs, our work also focuses on the precise evaluation of LLMs, motivated by the need of improving diagnostic performance and ensuring diagnostic safety (Balogh et al., 2015; Donaldson et al., 2000). Existing evaluation metrics for LLM output are insufficient for evaluating diagnostic accuracies, where precise performance is necessary to ensure diagnostic safety. We focus on an evaluation framework that can identify the diagnostic errors and the root cause, and assess the self-explanatory aspects of LLMs’ diagnostic processes. We designed the first human evaluation survey, following the SaferDX instrument, an organizational self-assessment tool with recommended practices aimed at improving diagnostic safety (Singh et al., 2019), for LLMs diagnosis prediction. The survey also incorporates the latest evaluation criteria for LLM, including factual accuracy, hallucination, quality of evidence, and other relevant aspects, identified from previous work in the field of biomedical NLP (Otmakhova et al., 2022; Singhal et al., 2023; Moramarco et al., 2021; Adams et al., 2021). Our aim is to bridge the gap between comprehensive diagnostic evaluation for safety and the capabilities of advanced language models, facilitating a deeper understanding of their diagnostic performance, and paving the way for safe LLM-augmented diagnostic decision support.

Our work and contribution are structured into four primary components:

1. designing and evaluating Dr.Knows, a graph-based model that selects the top N probable diagnoses with explainable paths (§4);
2. designing and implementing the first human evaluation framework for LLMs diagnosis generation and reasoning (§4.3),
3. revealing the usefulness of Dr.Knows as an additional module to augment LLMs in generating relevant diagnoses, a first iteration of integrating knowledge graphs for graph prompting (§2.3),
4. demonstrating the utilities of our proposed human evaluation framework that reveals LLM’s diagnostic performance with critical aspects of ensuring diagnostic safety (§2.3).

Our research poses a new problem that has not been addressed in the realm of NLP for diagnosis generation - harnessing the power of knowledge graphs for the controllability and explainability of LLMs. The following key findings will inform future work on developing knowledge graph-based methods for LLMs for diagnostic prediction:

### Strong Diagnostic Performance

Using the proposed human evaluation framework, ChatGPT demonstrated robust diagnostic accuracy with a median score of 66%, supported by exceptional self-explanation capabilities (“Reasoning” median score > 94%), underscoring its potential as a clinical diagnostic decision-support tool.

### Knowledge Graph’s Impact on Abstraction and Correct Reasoning

Integrating knowledge graphs into ChatGPT had a notable impact on finding the correct medical concepts, enhancing the model’s ability to generate abstractive diagnoses and improving reasoning (rationale sub-category in human evaluation) by guiding the diagnostic process with relevant knowledge paths.

### Future Knowledge Graph Model Enhancements

Analysis of Dr.Knows highlighted limitations in cases with unrelated pathways. Addressing these challenges through improving clinical narrative embedding, as well as improving the design of Dr.Knows with other components like Bayesian network, might enhance the diagnostic potential of KG-based models in the future.

### Utility of Our Proposed Human Evaluation for LLM

While the overall diagnostic accuracy and reasoning scores show the output with and without knowledge paths in the input has no differences, the broken-down scores present the strengths and weaknesses of different models. The granular approach of evaluation enables a more informed analysis of LLMs for particular applications and contributes to the iterative process of model refinement. The scoring aspects of plausibility, omission, specificity, rationale address various critical facets of AI interpretability and decision-making quality, aiming at mitigating the risks and enhancing the reliability and safety of diagnostics provided by AI systems. We provide the full guidelines of human evaluation in Supplementary Material and hope to contribute to facilitating the development of safe AI diagnostic tools.

Figure 1 presents the study overview of this work. We studied summarizing diagnoses from daily progress notes written in the SOAP-format. We developed a novel graph model, Dr.Knows, that identifies and retrieves relevant knowledge paths from UMLS KG. Dr.Knows is available in two versions: *TriAttn*_*w*_, which employs trilinear attention to determine the relevance scores for each knowledge path, and *MultiAttn*_*w*_, which utilizes a multi-head attention mechanism to score and select knowledge paths. Our initial evaluation of Dr.Knows focuses on its capability to identify and predict Concept Unique Identifiers (CUIs) for diagnoses, specifically addressing the *CUI prediction task*. Subsequently, we explored how these additional knowledge pathways could be harnessed to enhance ChatGPT’s ability to summarize diagnoses derived from daily progress notes. To achieve this, we integrated the knowledge pathways predicted by Dr.Knows into a prompting framework for ChatGPT. Additionally, we presented the performance difference between the zero-shot and few-shot settings.

**Figure 1:**
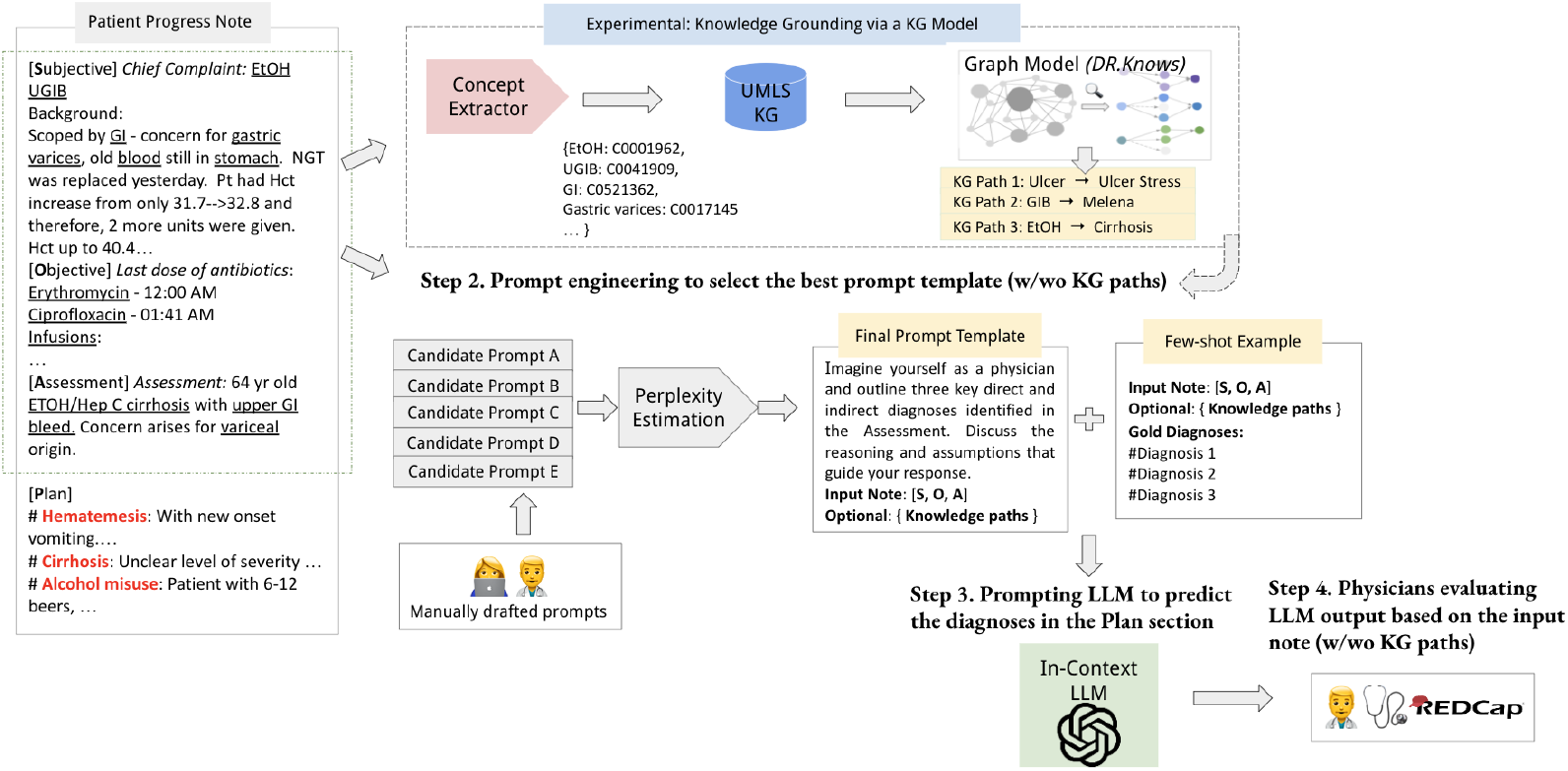
Study overview: we focused on generating diagnoses (red font text in the Plan section) using the SOAP-Format Progress Note with the aid of large language models (LLM). The input consists of Subjective, Objective and Assessment sections (the dotted line box on the example progress note), and diagnoses in the Plan sections are the ground truth. We introduced an innovative knowledge graph model, namely DR.KNOWS, that identifies and extracts the most relevant knowledge trajectories from the UMLS Knowledge Graph. The nodes for the UMLS knowledge graph are Concept Unique Identifiers (CUIs) and edges are the semantic relations among CUIs. We experimented with prompting ChatGPT for diagnosis generation, with and without DR.KNOWS predicted knowledge paths. Furthermore, we investigated how this knowledge grounding influences the diagnostic output of LLMs using human evaluation. Text with underlines are the UMLS concepts identified through a concept extractor.

**Figure 2:**
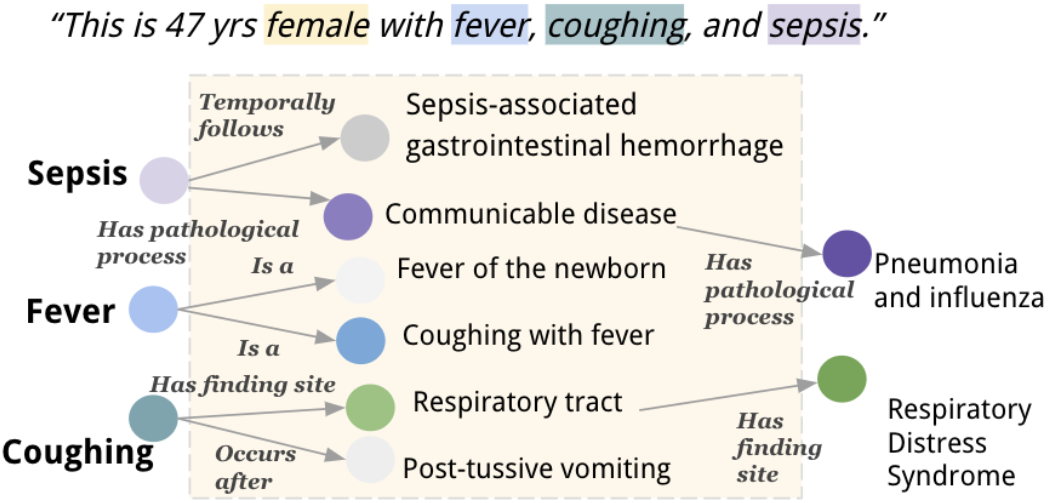
Inferring possible diagnoses within 2-hops from a UMLS knowledge graph given a patient’s medical description. We highlight the UMLS medical concept in the color boxes (“female”, “sepsis”, etc). Each concept has its own subgraph, where concepts are the vertices, and semantic relations are the edges (for space constraint, we neglect the subgraph for “female” in this graph presentation). On the first hop, we could identify the most relevant neighbor concepts to the input description. The darker color the vertices are, the more relevant they are to the input description. A second hop could be further performed based on the most relevant nodes, and reach the final diagnoses “Pneumonia and influenza” and “Respiratory Distress Syndrome”. Note that we use the preferred text of Concept Unique Identifiers (CUIs) for presentation purposes. The actual UMLS KG is built on CUIs rather than preferred text.

We summarized the deployment of evaluation metrics in Table 2. On CUI prediction task, we reported CUI-based Recall, Precision and F-score. The metrics helped us understand if the Dr.Knows could accurately identify CUIs that are the final diagnoses. On the results obtained through ChatGPT, we first applied automated metrics including CUI-based Recall, Precision, F-score, and two ROUGE variants (ROUGE-2 and ROUGE-L Lin (2004)). Then we asked two medical professionals to conduct a human evaluation using our proposed framework under the supervision of two senior physicians. By examining the effects of graph prompting on LLMs with real-world EHR data, we strive to contribute to an explainable AI diagnostic pathway.

## 2 Results

### 2.1 Data overview

We used two sets of progress notes from different clinical settings in this study: mimic-iii and in-house EHR datasets. mimic-iii is one of the largest publicly available databases that contains de-identified health data from patients admitted to intensive care units (ICUs), developed by the Massachusetts Institute of Technology and Beth Israel Deaconess Medical Center (BIDMC). mimic-iii includes data from over 38,000 patients admitted to ICUs at the BIDMC between 2001 and 2012. The second set, namely the in-house EHR data, was a subset of EHRs including adult patients (ages > 18) admitted to the Univesity of Wisconsin Health System between 2008 to 2021. In contrast to the mimic subset, the in-house set covered progress notes from all hospital settings, including Emergency Department, General Medicine Wards, Subspecialty Wards, etc. While the two datasets originated from separate hospitals and departmental settings and might reflect distinct note-taking practices, they both followed the SOAP documentation format for progress notes.

Gao et al. (2022, 2023) introduced a subset of 1005 progress notes from mimic-iii with active diagnoses annotated from the Plan sections. Therefore, we applied this dataset for training and evaluation for both graph model intrinsic evaluation (§2.2) and diagnosis summarization (§2.3). The in-house dataset did not contain human annotation. Still, by parsing the text with a medical concept extractor that was based on UMLS SNOMED-CT vocabulary, we were able to pull out concepts that belonged to the semantic type of T047 Disease and Syndromes. We deployed this set of concepts as the ground truth data to train and evaluate the graph model in §2.2. The final set of in-house data contained 4815 progress notes. We presented the descriptive statistics in Table 1. When contrasting with mimic-iii, the in-house dataset exhibited a greater number of CUIs in its input, leading to an extended CUI output. Additionally, mimic-iii encompassed a wider range of abstractive concepts compared to the progress notes of in-house. Example Plan sections from the two datasets are in the Appendix A.

**Table 1:** Average number of unique Concept Unique Identifiers (CUI) in the input and output on the two EHR dataset: MIMIC-III and IN-HOUSE. Abstractive concepts are those not found in the input, but present in the gold standard diagnoses.

| Dataset | Dept. | #Input CUIs | #Output CUIs | % Abstractive Concepts |
| --- | --- | --- | --- | --- |
| MIMIC-III | ICU | 15.95 | 3.51 | 48.92% |
| IN-HOUSE | All | 41.43 | 5.81 | <1% |

**Table 2:** Overview of the evaluation for the tasks. Note that for CUI-based evaluation, the text is first converted to a set of UMLS CUIs using a concept extractor.

| Evaluation metrics | Description | Tasks |
| --- | --- | --- |
| CUI-based Recall, Precision, F-score | An automated metric measuring the set overlap of predicted CUIs and gold CUIs. Formulas are described in §4 | CUI prediction (§2.2);<br>Diagnosis prediction (§2.3) |
| ROUGE-2, ROUGE-L<br>Lin (2004) | Automated metrics measuring the overlap of the bigrams (ROUGE-2) or longest common sub-sequences (ROUGE-L) between predicted text and reference text. | Diagnosis prediction (§2.3) |
| Human evaluation | Medical professionals evaluating ChatGPT’s diagnosis output and reasoning output. Sub-categories include accuracy, omission, abstraction, plausibility, specificity and others. Details can be found at §4.3 | Diagnosis prediction (§2.3) |

Given that our work encompasses a public EHR dataset (mimic-iii) and a private EHR dataset with protected health information (in-house), we conducted training using three distinct computing environments. Specifically, most of the experiments on mimic-iii were done on Google Cloud Computing (GCP), utilizing 1-2 NVIDIA A100 40GB GPUs, and a conventional server equipped with 1 RTX 3090 Ti 24GB GPU. The in-house EHR dataset is stored on a workstation located within a hospital research lab. The workstation operates within a HIPAA-compliant network, ensuring the confidentiality, integrity, and availability of electronic protected health information (ePHI), and is equipped with a single NVIDIA V100 32GB GPU. To use ChatGPT, we utilized an in-house ChatGPT-3.5-turbo version hosted on our local cloud infrastructure. This setup ensures that no data is transmitted to OpenAI or external websites, and we are in strict compliance with the MIMIC data usage agreement.

### 2.2 Evaluation of DR.KNOWS on Predicting Diagnoses

We compared Dr.Knows with QuickUMLS (Soldaini and Goharian, 2016), which is a concept extraction baseline that identified the medical concepts from raw text. We took input text, parsed it with the QuickUMLS and outputted a list of concepts. Table 3 provided results on the two EHR datasets mimic and in-house. The selection of different top *N* values was determined by the disparity in length between the two datasets (see App. A). Dr.Knows demonstrated superior precision and F-score across both datasets compared to the baseline, with precision scores of 19.10 (95% CI: 17.82 - 20.37) versus 13.59 (95% CI: 12.32 - 14.88) on MIMIC, and 22.88 (95% CI: 20.92 - 24.85) versus 12.38 (95% CI: 11.09 - 13.66) on the in-house dataset. Additionally, its F-scores of 25.20 on MIMIC and 25.70 on the in-house dataset exceeded the comparison scores of 21.13 (95% CI: 19.85 - 22.41) and 20.09 (95% CI: 18.81 - 21.37), respectively, underscoring its effectiveness in accurately predicting diagnostic CUIs. The TriAttn_*w*_ variant of Dr. Knows consistently outperformed the MultiAttn_*w*_ variant on both datasets, with F-scores of 25.20 (95% CI: 23.93 - 26.48) versus 23.10 (95% CI: 21.83 - 24.39) on MIMIC and 25.70 (95% CI: 24.06 - 27.37) versus 17.69 (95% CI: 16.40 - 18.96) on in-house. The concept extractor baseline reached the highest recall, with 56.91 on mimic and 90.11 on in-house, as it found all the input concepts that overlapped with the reference CUIs, in particular on the in-house dataset that was largely an extractive dataset (App. A).

**Table 3:**
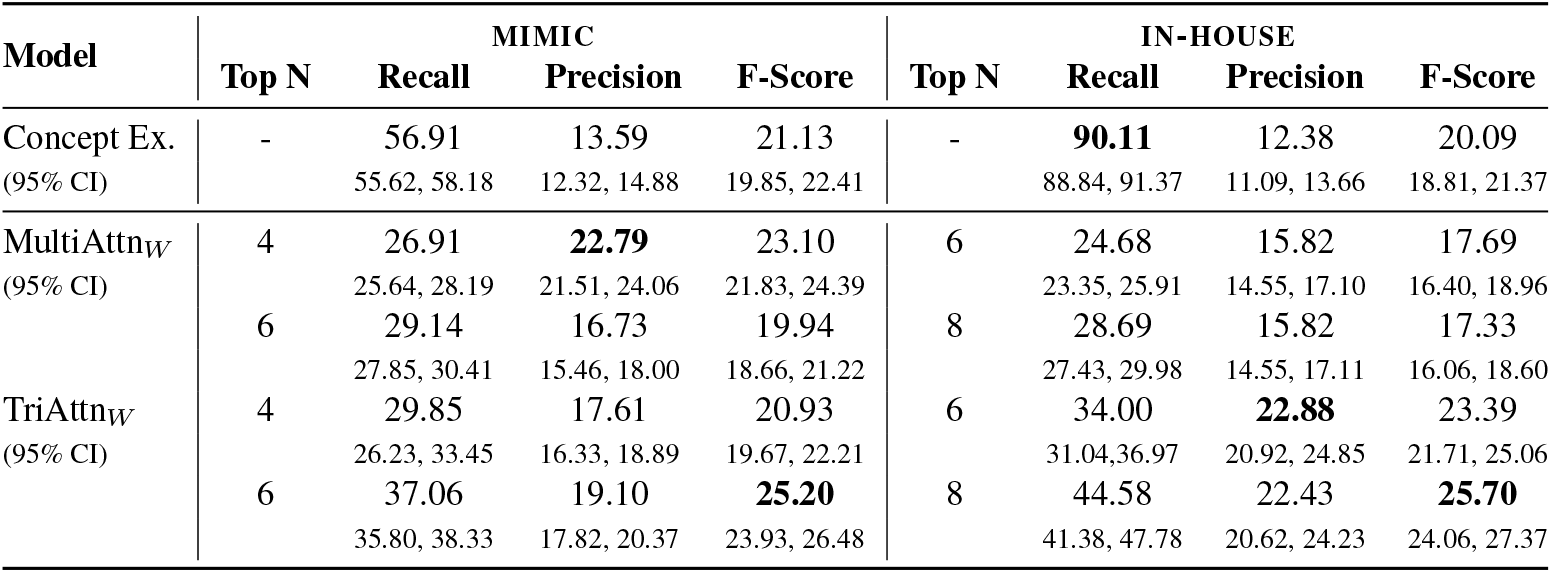
Performance comparison between concept extraction (Concept Ex.) and two DR.KNOWS variants on target CUI prediction using MIMIC and IN-HOUSE dataset.

### 2.3 Prompting Large Language Models for Diagnosis Generation

#### Results reported in automated metrics

Shifting from a zero-shot to a few-shot learning scenario resulted in a clear boost in performance. The few-shot’s minimum ROUGE-2 score of 9.63 (95% CI: 8.32 - 10.06), surpassed the zero-shot’s maximum of 7.05 (95% CI: 6.54 - 7.56), and the few-shot’s minimum CUI-F score of 20.96 (95% CI: 20.19 - 21.73) outperformed zero-shot’s score of 18.21 (95% CI: 17.46. - 18.98).

The performance comparison between ChatGPT with Dr.Knows in the predicted paths scenario versus the no paths scenario provided additional improvement in the few-shot setting. Notably, in the 3-shot scenario, the +Path yielded a ROUGE-L score of 24.32 (95% CI: 22.44 - 24.25) versus 21.84 (95% CI: 19.99 - 22.09), and a CUI-F score of 25.30 (95% CI: 24.52 - 26.06) versus 21.02 (95% CI: 20.26 - 21.79) from the no path scenarios. In the 5-shot setting, the +Path configuration outperformed the no path setting across all metrics (Table 4). The ROUGE-2 score was 11.73 (95% CI: 10.51 - 12.25), and exceeded the no path score of 9.73 (95% CI: 8.52 - 10.18). ROUGE-L scores were also higher at 25.43 (95% CI: 23.53 - 25.35) compared to 21.23 (95% CI: 19.58 - 21.71).

**Table 4:**
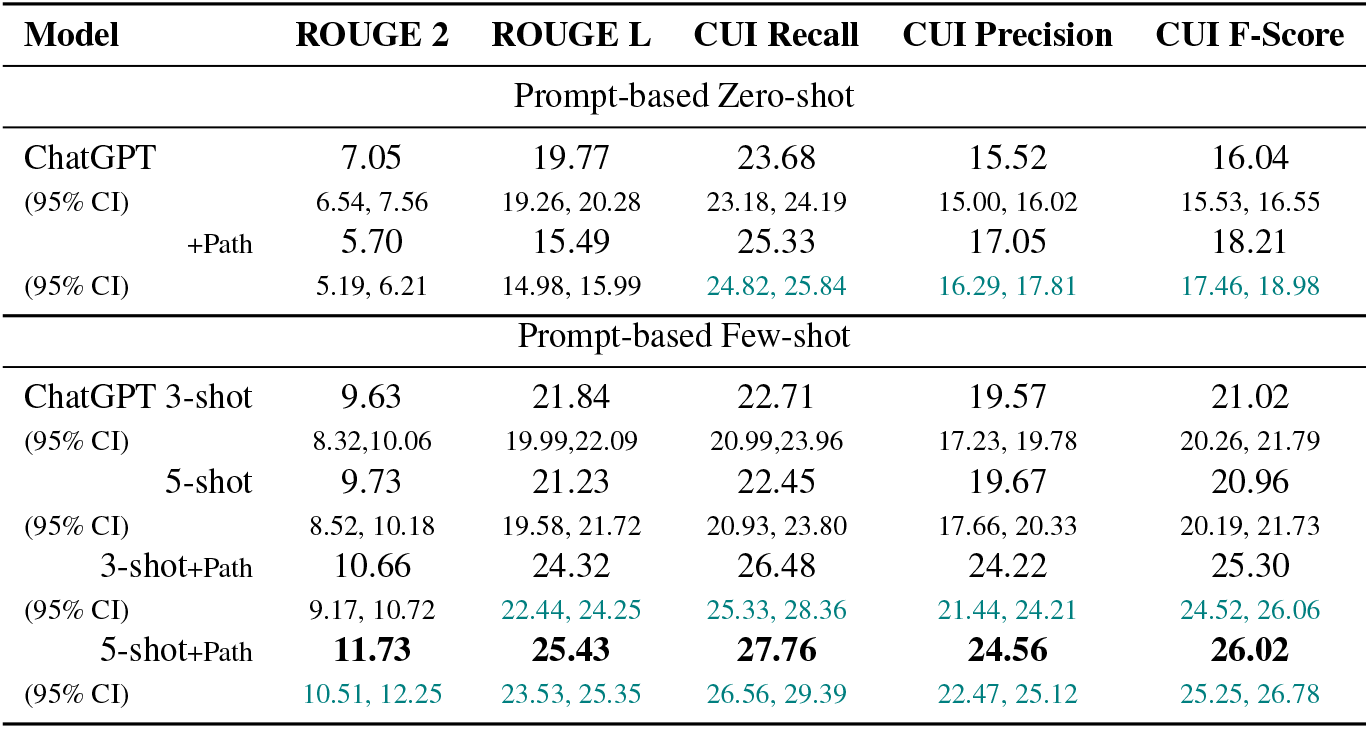
Best performance on MIMIC test set (with annotated active diagnoses) from ChatGPT across all prompt styles with (+Path) and without DR.KNOWS path prompting. We report ROUGE-2, ROUGE-L, CUI Recall, Precision and F-score to illustrate the performance difference better. We use teal color to highlight the 95% confidence interval (CIs) when there is a distinct CIs for the +Path compared to no path scenarios.

#### Results from human evaluation

Human evaluation was performed on few-shot ChatGPT with and without KG, using 38.88% samples of the test set (n=92). Figure 3 shows the diagnosis scores and reasoning scores from ChatGPT with and without Dr.Knows. Both models achieved diagnostic accuracy with a median score surpassing 0.66 (IQR: 0.57-0.74 for with knowledge graph (KG); IQR 0.54-0.75 for no KG), and their reasoning scores exhibited a median exceeding 0.90 (IQR: 0.86-0.97 for KG; 0.90-0.97 for no KG). In contrast to the automated metrics, human evaluation indicated that the presence or absence of Dr.Knows did not yield an overall difference in performance (p=0.63); however, several subgroup components were different.

**Figure 3:**
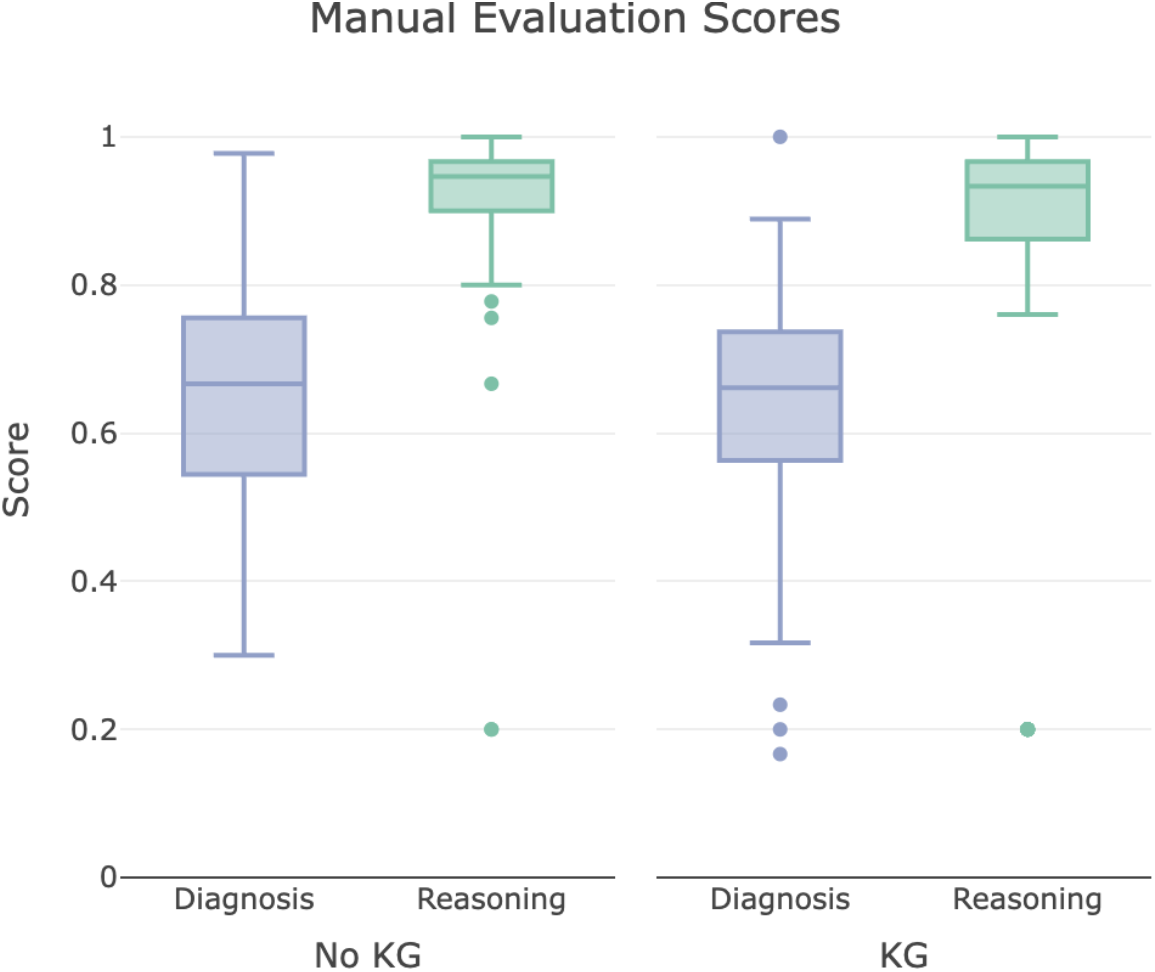
Overall performance for ChatGPT models with the absence (“No KG”) and the presence of Dr.Knows (“KG”).

Figure 4 describes all components of the diagnosis scores, considering six distinct scoring aspects. ChatGPT models with and without KG paths exhibited similar performance in accuracy, omission, uncertainty, plausibility, and specificity. Notably, both models excelled in terms of accuracy, consistently providing about 80% affirmative answers (“Yes”) to the question of whether the output meets the criteria for an official diagnosis. In contrast, their performance in abstraction ranged from 13% (“KG”) to 18% (“No KG”). On omitted diagnoses, approximately 14% to 15% stemmed from aleatoric uncertainty. This uncertainty contributed to about 18% of cases for “majority aleatoric” and 33% for “all aleatoric” scenarios for both models. Lastly, concerning the level of abstraction, ChatGPT with KG did not favor more extractive diagnoses than ChatGPT without KG, scoring 87% compared to 81% for “No” answers (p=0.09).

**Figure 4:**
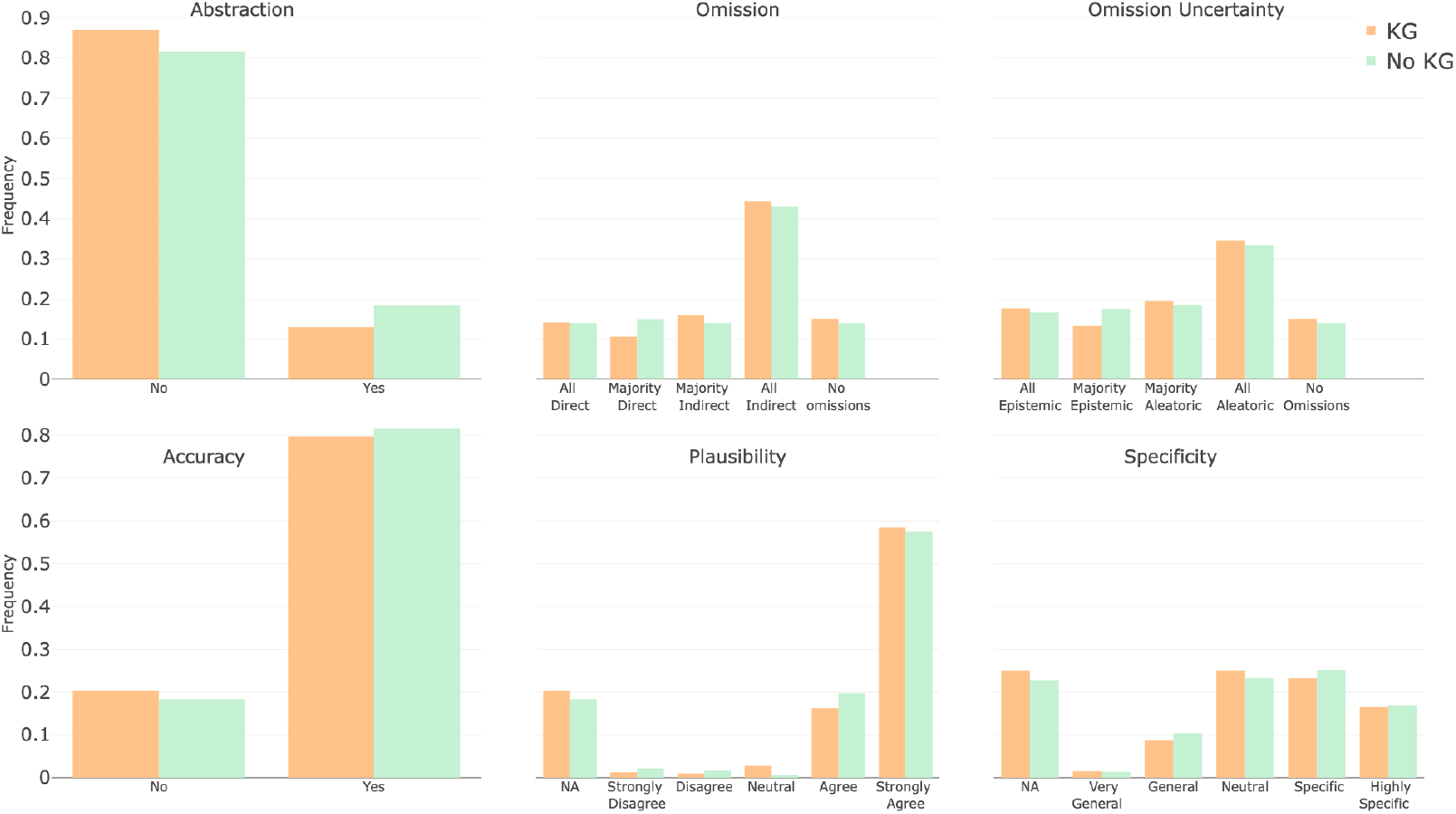
Diagnosis scores for ChatGPT models with the absence (“No KG”) and the presence of Dr.Knows (“KG”).

In Figure 5, when examining the reasoning scores, there was no significant increase in omission, with 16% observed with KG, as opposed to 10% without KG (p=0.16). When it comes to rationale (correct reasoning), ChatGPT with KG exhibits a 55% strong agreement with humans, while ChatGPT “No KG” demonstrates 50% strong agreement (p<0.01). On the abstraction category asking about the presence of abstraction in model output, there was a notable drop from 88% (“No KG”) to 78% (“KG”) in the affirmative responses (p=0.03), indicating less abstraction required with KG paths. Differences were also noted in effective abstraction in favor of the KG paths (p<0.01).

**Figure 5:**
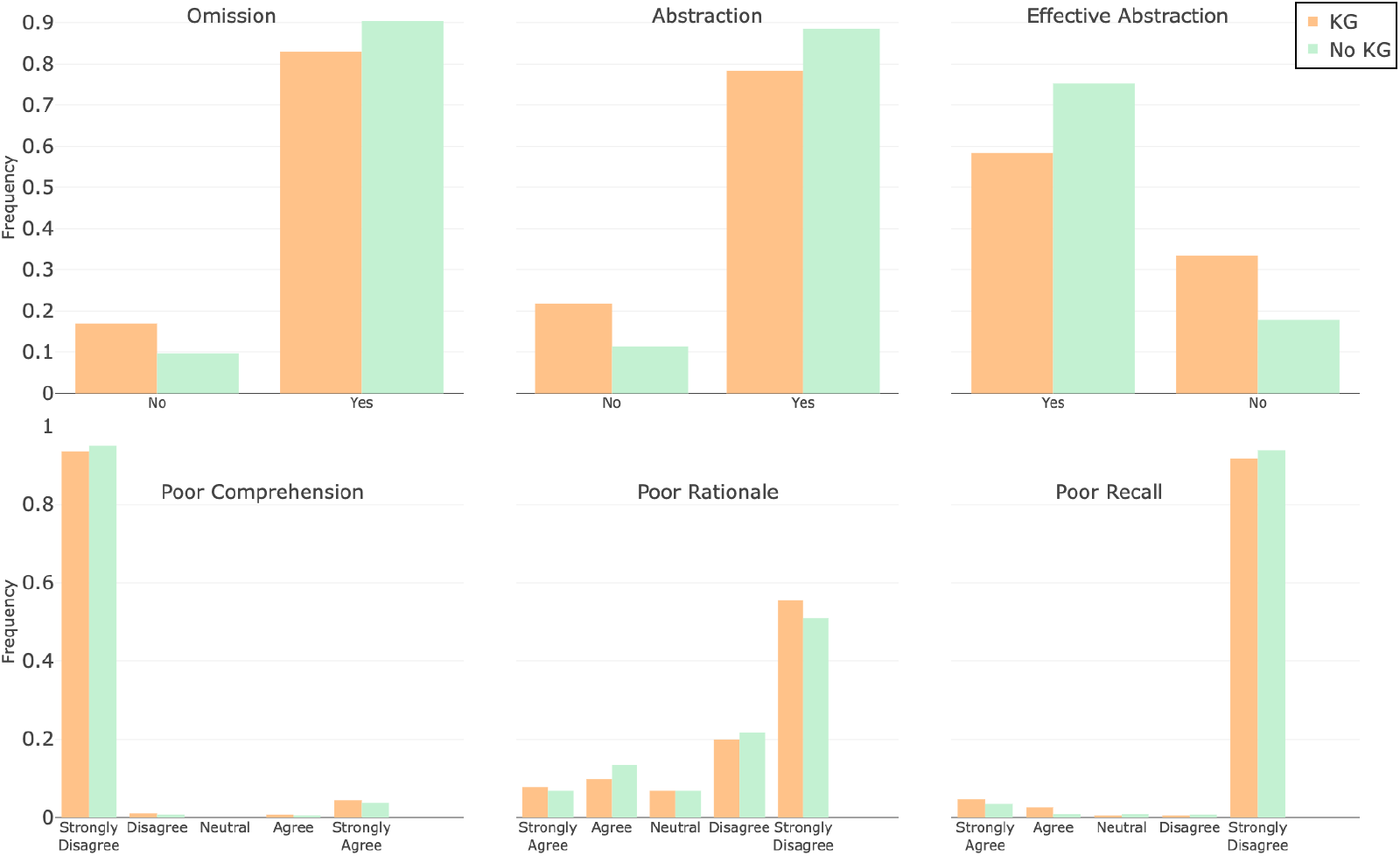
Reasoning scores for ChatGPT models with the absence (“No KG”) and the presence of Dr.Knows (“KG”).

#### Error analysis

We discovered two primary types of error in Dr.Knows output that could result in missed opportunities for improving knowledge grounding. Figure 6 presents an example where the ChatGPT did not find the provided knowledge paths useful. In this case, the majority of the provided knowledge paths were highly extractive (“leukocytosis” “reticular dysgenesis”“paraplegia” are target concepts the knowledge paths led to and all have “self” relationship). On the abstraction paths the target concepts “abdomen hernia scrotal” and “chronic neutrophilia” were found, which were not relevant to the input patient condition.

**Figure 6:**
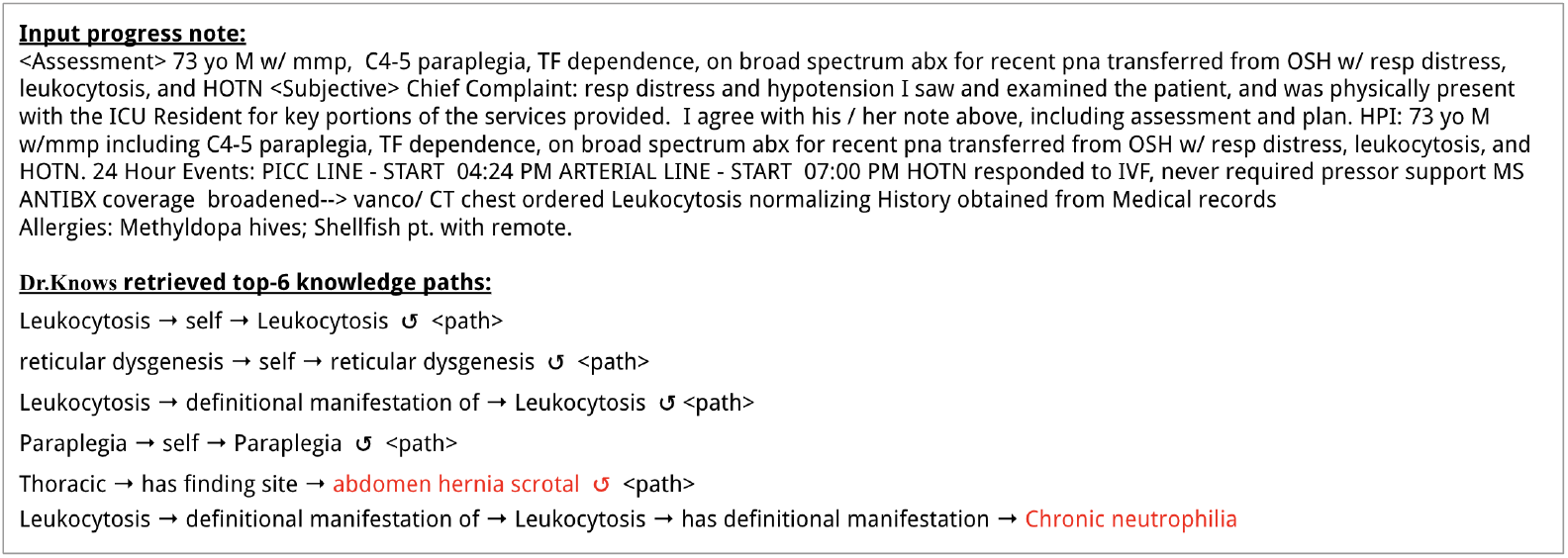
An error example of Dr.Knows retrieved knowledge pathways. Dr.Knows finds two paths leading to irrelevant and misleading diagnosis, marked as red fonts. The ↺ symbol represents a self-loop.

Another error observed occurred when Dr.Knows selected the source CUIs that were less likely to generate pertinent pathways for clinical diagnoses, resulting in ineffective knowledge paths. Figure 7 shows a retrieved path from “Consulting with (procedure)” to “Consultation-action (qualifier value)”. Although some procedure-related concepts like endoscopy or blood testing were valuable for clinical diagnosis, this specific path of consulting did not contribute meaningfully to the input case. Similarly, another erroneous pathway began with “Drug Allergies” and led to “Allergy to dimetindene (finding)”, which is contradictory given that the input note explicitly states “No Known Drug Allergies”. While the consulting path’s issue was its lack of utility, the “Drug Allergies” path could introduce the risk of hallucination (misleading or fabricated content) within ChatGPT.

**Figure 7:**
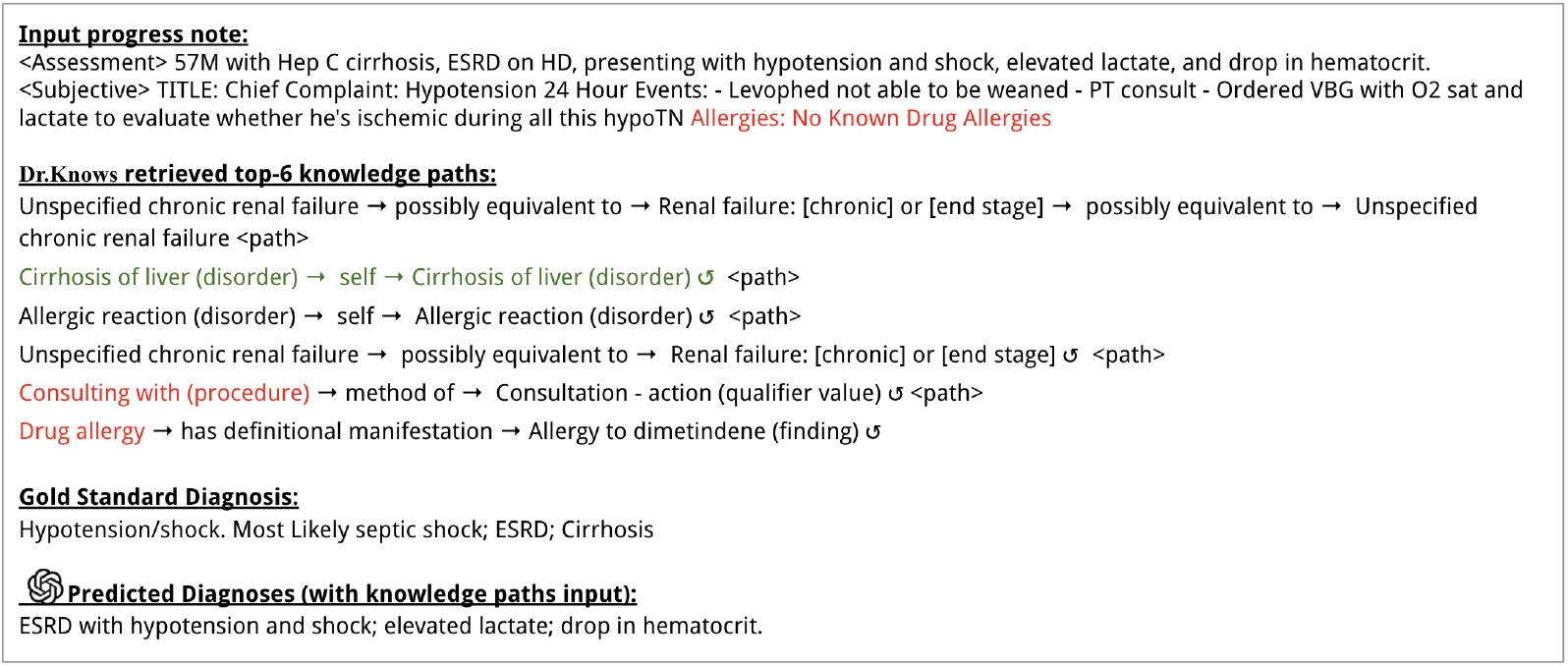
An example from ChatGPT with Dr.Knows extracted knowledge pathways. Two paths had source CUIs (“Consulting with (procedure), Drug Allergy”) that were less likely to generate pertinent paths for clinical diagnoses. Note that the path of “Drug allergy” led to a path contradicting to the “No Known Drug Allergies” description in the input. The path of “cirrhosis of liver” was a correct diagnosis, but ChatGPT failed to include it.

In addition to Dr.Knows’ errors, there were instances where ChatGPT failed to leverage accurate knowledge paths presented. Figure 7 includes a knowledge path about “Cirrhosis of liver”, which was a correct diagnosis. However, ChatGPT did not contain this diagnosis.

Finally, when Dr.Knows retrieved the correct knowledge paths and ChatGPT utilized it well, there was an improvement in the output quality. Figure 8 presents an example where all the paths retrieved by Dr.Knows were relevant to the input, and successfully led to ChatGPT outputting plausible diagnoses. This led to higher plausibility scores from human evaluators.

**Figure 8:**
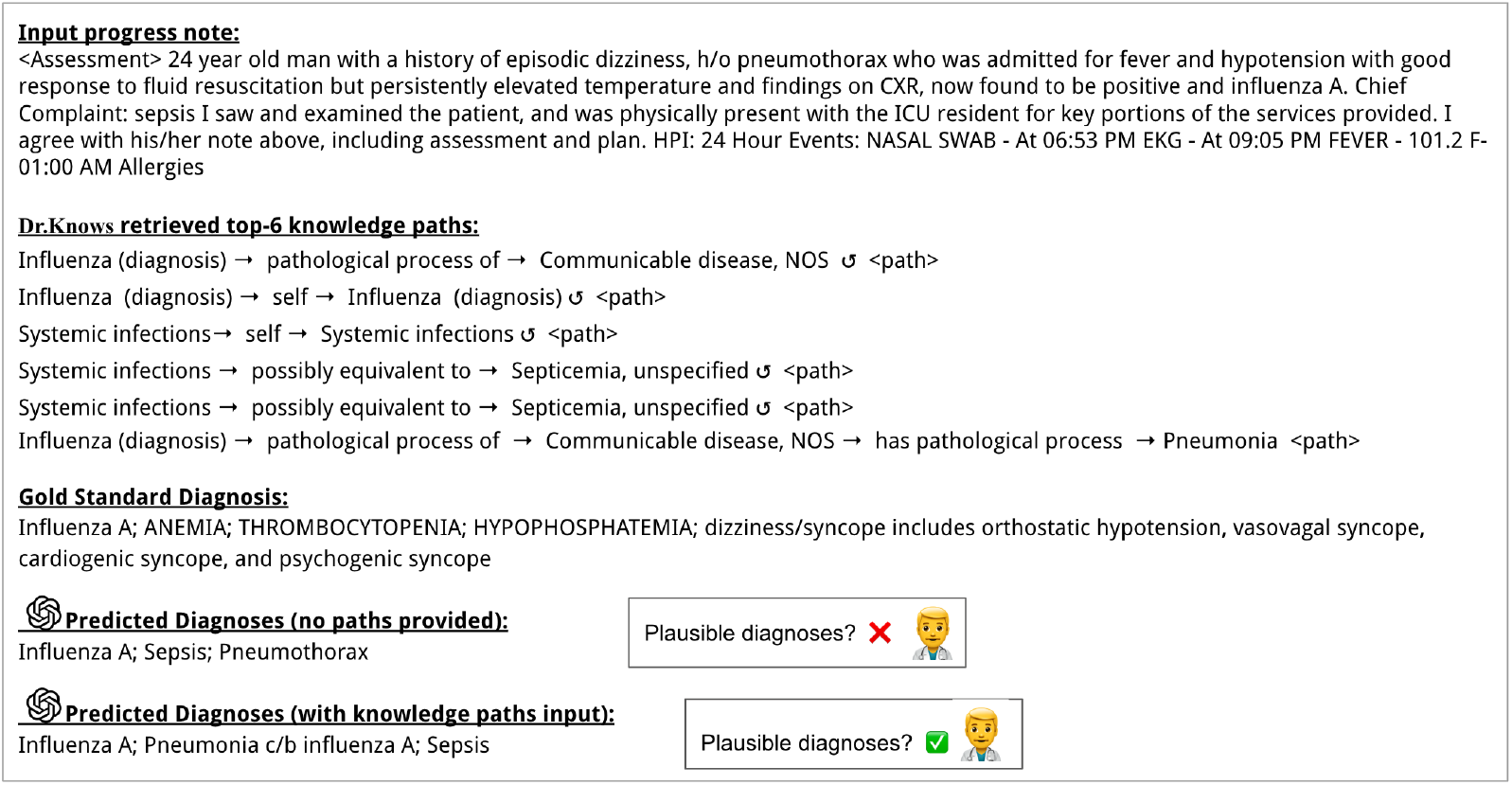
Illustration of ChatGPT with KG paths selected by Dr.Knows, introducing abstraction concepts and enhancing predicted diagnosis plausibility. Human annotators find the predicted diagnosis from ChatGPT, with knowledge paths, more plausible compared to the one without.

**Figure 9:**
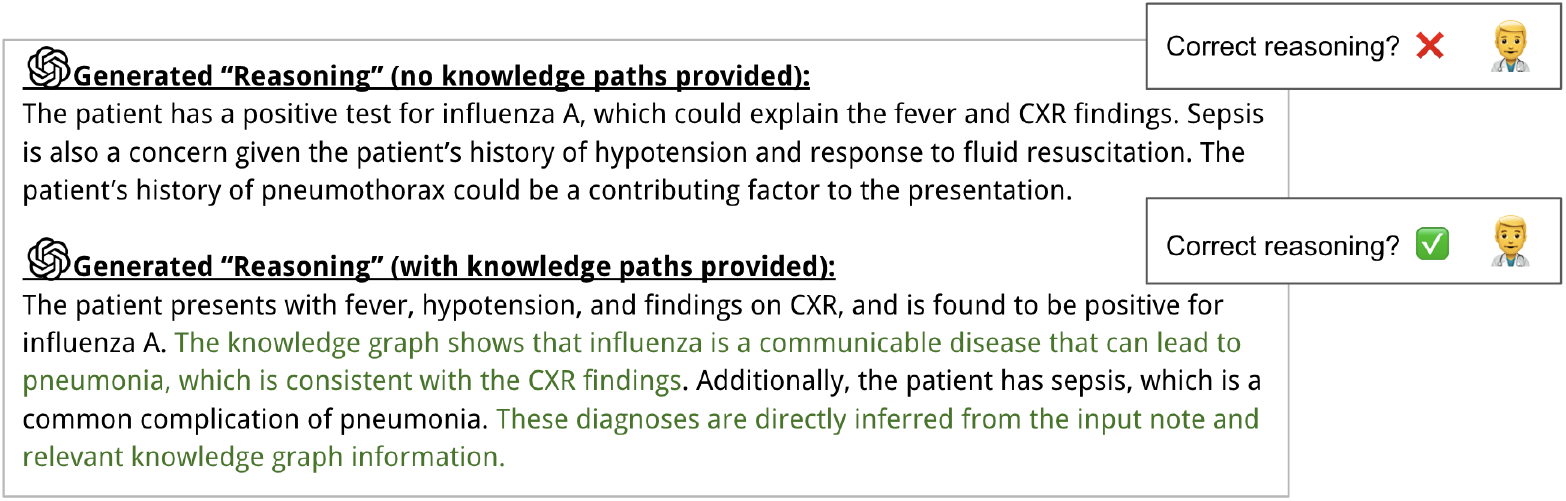
The “Reasoning” output of ChatGPT with and without knowledge paths on the same input example as in Fig 8. The highlighted green text demonstrates the utilization of Dr.Knows knowledge paths to enhance ChatGPT’s reasoning capabilities. Human evaluators find that the ChatGPT with KG produces correct reasoning.

## 3 Discussion

On the few-shot setting, with and without Dr.Knows retrieved paths, ChatGPT demonstrated a median diagnostic accuracy of 66% and exhibited a remarkable median score exceeding 94% in reasoning, as per human evaluation. The incorporation of Dr.Knows retrieved paths proved to be beneficial, enhancing ChatGPT’s performance, as evidenced by higher scores from automated metrics and improvements noted in abstraction and rationale aspects during human evaluation. A primary source of errors stemmed from Dr.Knows incorrectly identifying irrelevant target concepts and initiating retrievals with less effective CUIs. This issue, along with ChatGPT’s struggle to incorporate the correct paths, was highlighted as key areas for improvements.

### Impact of KG on LLM knowledge grounding

Based on human evaluation of overall diagnostic accuracy and reasoning, integrating a knowledge graph appeared to make no noticeable impact on the performance of ChatGPT. However, closer examination of the scoring sub-category revealed that Dr.Knows enhances ChatGPT’s ability to identify abstractive diagnoses and accurately deduce connections between input and possible diagnoses. Half diagnoses within MIMIC dataset are not abstracted (< 50%), which may have limited the ability of the knowledge graph approach to demonstrate benefits over the native LLM, as the knowledge graph approach would be expected to specifically augment the abstraction task. The 10% decrease in ChatGPT’s abstraction with KG can be attributed to the more abstract information provided in the input when using KG. Human evaluation also favored ChatGPT with KG’s rationale rather than without KG (p<0.01), indicating that the inclusion of KG enhances the medical grounding of the responses, leading to more clinically relevant and factually supported reasoning. Results evaluated by automated metrics, ROUGE and Concept F-score also illustrated the improved precision and F-score in identifying the correct diagnostic concepts. Such knowledge grounding highlighted the potential for strengthening LLM’s medical decision-making and reducing hallucinations, which is critical in an AI-augmented diagnostic decision-support system.

Through these results, our work presented the potential benefits of knowledge grounding through a retrieval-augmented generation framework utilizing the most important concepts and relations for knowledge-intensive tasks. Expanding or modifying the memory and knowledge of large language models is not a straightforward task, potentially resulting in factual inaccuracies and hallucinations. The use of a retrieve-and-augment framework, leveraging external knowledge sources, has demonstrated its ability to mitigate these issues, as evidenced by previous research (Lewis et al., 2020; Shuster et al., 2021).

### Overall performance and insights drawn from human evaluation scores

The median diagnostic accuracy of 66%, achieved by both few-shot prompting ChatGPT models, revealed ChatGPT’s robust performance in generating diagnoses from daily hospital progress notes. The exceptionally strong performance in reasoning, with a median score surpassing 94%, highlights ChatGPT’s capacity for weighing and integrating various pieces of evidence when arriving at a diagnosis, a promising indication for clinical diagnostic reasoning. This evidence-based approach is crucial for LLMs for clinical diagnostic decision-support, ensuring that the model’s recommendations are rooted in the provided input and that such evidence-based grounding is accessible to healthcare providers.

The detailed scoring in human evaluation not only highlighted ChatGPT’s performance but also pointed towards areas for future enhancement. One significant issue to address was the omission of diagnoses. Currently, ChatGPT exhibited no omitted diagnoses in only 15% of cases, with the majority of omitted diagnoses attributed to aleatoric uncertainty. This uncertainty arises when the evidence for diagnoses is present in the input, but the model fails to accurately capture and incorporate this information. Addressing and minimizing this type of uncertainty is pivotal for enhancing the precision and reliability of the diagnostic process using ChatGPT.

### Discrepancy between automated metrics and human evaluation

Our experiments revealed intriguing differences between the results obtained from automated metrics and human evaluation. While the automated metrics suggest a performance difference between the two models, with the KG-augmented model demonstrating a performance gain over its non-KG counterpart, human evaluation results show that both models are consistently rated as equally proficient. We attributed this divergence to the specific dimension assessed by automated metrics, as opposed to human evaluation scores that aggregate multiple distinct scoring criteria. ROUGE assesses content quality through string overlap analysis, while the concept-based F-score gauges the precision of identified concepts in the generated text. These metrics offer distinct perspectives on model performance. Nevertheless, it is important to recognize that these metrics may not entirely capture the nuanced aspects of human evaluation. Further investigation on the correlation between automated metrics and human scoring is concluded as future work. We also encourage future research to explore ways to bridge the gap between automated metrics and human judgment for a more comprehensive assessment of model performance.

### Informing future knowledge graph model development from Dr.Knows error analysis

Error analysis showed that Dr.Knows still suffered from recognizing knowledge paths that were not related to the input patient representation, and that the selection of starting medical concepts was pivotal in finding the right paths. Currently, Dr.Knows relied solely on semantic-based ranking on the candidate paths, that is, the cosine similarity between candidate path embeddings and input text, with the quality of these embeddings being crucial for ranking performance. In addition to enhancing the representation method and these embeddings, other elements that are essential in modeling relations between symptoms and diagnoses, for instance, probabilistic modeling (Rotmensch et al., 2017; Wan and Du,2021), should be incorporated into the graph-based methods. We encourage future research to explore this integration and improve Dr. Knows’ diagnostic potential.

The error analysis also presented instances where ChatGPT neglected to incorporate certain beneficial knowledge paths. It’s important to acknowledge that ChatGPT operates as a black-box API model, with its internal weights and training processes being inaccessible. To enhance the efficacy of the graph-based retrieve-and-augment framework, it would be advantageous to explore the potential of graph-prompting and instruction tuning on open-source language models. These methods could refine the model’s ability to utilize relevant information effectively. Other relevant research also employs advanced prompting techniques, such as self-retrieval-augmented generation (Asai et al., 2023) and step-back prompting (Zheng et al., 2023), which merit further exploration in future investigations.

In conclusion, LLMs like ChatGPT are a promising direction for generating diagnoses for clinical decision support; however, methods such as graph prompting are needed to guide the model down correct reasoning paths to avoid hallucinations and provide comprehensive diagnoses. While we show some progress in a graph prompting approach with Dr.Knows, more work is needed to improve methods that leverage the UMLS knowledge source for grounding to achieve more accurate outputs. Furthermore, our human evaluation framework carries strong face validity and reliability to evaluate a model’s strengths and weaknesses as a diagnostic decision support system.

## 4 Methods

### 4.1 Grounding Medical Knowledge with Knowledge Graph

#### 4.1.1 Problem Formulation

##### Diagnosis in progress notes

Daily progress notes are formatted using the SOAP Format (Weed, 1969). The *Subjective* section of a SOAP format daily progress note comprises the patient’s self-reported symptoms, concerns, and medical history. The *Objective* section consists of structural data collected by healthcare providers during observation or examination, such as vital signs (e.g., blood pressure, heart rate), laboratory results, or physical exam findings. The *Assessment* section summarizes the patient’s overall condition with a focus on the most active problems/diagnoses for that day. Finally, the *Plan* section contains multiple subsections, each outlining a diagnosis/problem and its treatment plan. Our task is to predict the list of problems and diagnoses that are part of the *Plan* section.

##### Using UMLS KG to find potential diagnoses given a patient’s medical narrative

The UMLS concepts vocabulary comprises over 187 sources. For our study, we focused on the Systematized Nomenclature of Medicine-Clinical Terms (SNOMED CT). The UMLS vocabulary is a comprehensive, multilingual health terminology and the US national standard for EHRs and health information exchange. Each UMLS medical concept is assigned a unique SNOMED concept identifier (CUI) from the clinical terminology system. We utilize semantic types, networks, and semantic relations from UMLS knowledge sources to categorize concepts based on shared attributes, enabling efficient exploration and supporting semantic understanding and knowledge discovery across various medical vocabularies.

Given a medical knowledge graph where vertices are concepts and edges are semantic relations, and an input text describing a patient’s problems, we could perform multi-hop reasoning over the graphs and infer the final diagnoses. Figure 1 demonstrated how UMLS semantic relations and concepts can be used to identify potential diagnoses from the evidence provided in a daily care note. The example patient presents with medical conditions of fever, coughing, and sepsis, which are the concepts recognized by medical concepts extractors (cTAKES (Savova et al., 2010) and QuickUMLS (Soldaini and Goharian, 2016)) and the starting concepts for multi-hop reasoning. Initially, we extracted the direct neighbors for these concepts. Relevant concepts that align with the patient’s descriptions were preferred. For precise diagnoses, we chose the top *N* most relevant nodes at each hop.

This section introduces the architecture design for Dr.Knows. As shown in Figure 10, all identified UMLS concepts with assigned CUI from the input patient text will be used to retrieve 1-hop subgraphs from the constructed large UMLS knowledge graph. These subgraphs are encoded as graph representations by a Stack Graph Isomorphism Network (SGIN) (Xu et al., 2019) and then fed to the Path Encoder, which generates path representations. The Path Ranker module assesses 1-hop paths by considering their semantic and logical association with the input text and concept, generating a score using the path representation, input text, and concept representation. The top N scores among the set of 1-hop neighbor nodes, aggregated from all paths pointing to those nodes, guide the subsequent hop exploration. In case a suitable diagnosis node is not found, termination is assigned to the self-loop pointing to the current node.

**Figure 10:**
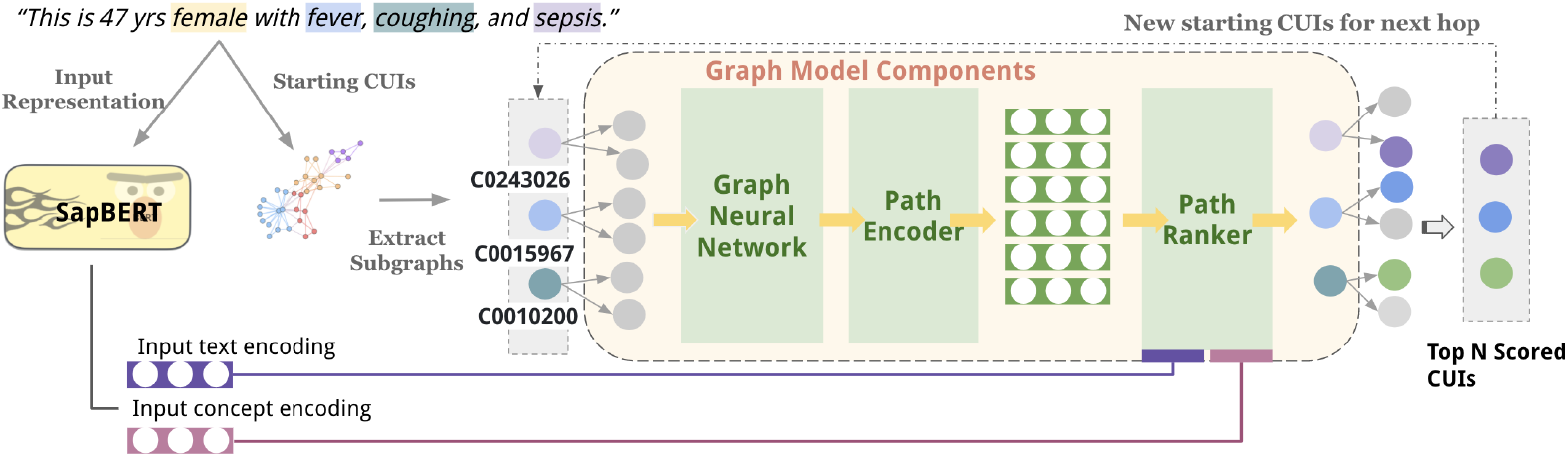
DR.KNOWS model architecture. The input concepts (“female”, “fever”, etc) are represented by concept unique identifiers (CUIs, represented as the combination of letters and numbers, e.g.”C0243026”, “C0015967”).

#### 4.1.2 Contextualized Node Representation

We defined the deterministic UMLS knowledge graph 𝒢 = 𝒱ℰ based on SNOMED CUIs and semantic relations, where 𝒱 is a set of CUIs, and ℰ is a set of semantic relations. Given an input text **x** containing a set of source CUIs 𝒱_*src*_ ⊆ 𝒱, and their 1-hop relations ℰ_*src*_ ⊆ ℰ, we can construct relation paths for each 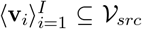 as **P** = {**p**_1_, **p**_2_, …, **p**_*J*_} s.t. **p**_*j*_ = {**v**_1_, **e**_1_, **v**_2_ … **e**_*t* 1_, **v**_*t*_}, *j* ∈ *J*, where t is a pre-defined scalar and *J* is non-deterministic. Relations **e**_*t*_ were encoded as one-hot embeddings. We concatenated all concept names for **v**_*i*_ with special token [SEP], s.t. **l**_*i*_ = [name 1 [SEP] name 2 [SEP] …], and encoded **l**_*i*_ using SapBERT (Liu et al., 2021) to obtain **h**_*i*_. This allowed the CUI representation to serve as the contextualized representation of its corresponding concept names. We chose SapBERT for its UMLS-trained biomedical concept representation. The **h**_*i*_ is further updated through topological representation using SGIN:

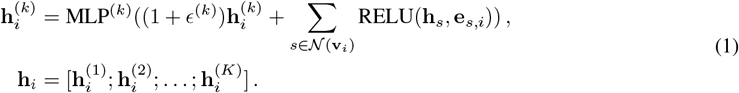

where 𝒩(**v**_*i*_) represents the neighborhood of node 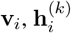 is the representation of node **v**_*i*_ at layer *k, ϵ*^(*k*)^ is a learnable parameter, and MLP^(*k*)^ is a multilayer perceptron. GIN iteratively aggregates neighborhood information using graph convolution followed by nonlinearity, modeling interactions among different **v** ⊆ 𝒱. Furthermore, the stacking mechanism is introduced to combine multiple GIN layers. The final node representation **v**_*i*_ at layer *K* is computed by stacking the GIN layers, where [*·*; *·*] denotes concatenation.

We empirically observed that some types of CUIs are less likely to lead to useful paths for diseases, e.g., the concept “recent” (CUI: C0332185) is a temporal concept and the neighbors associated with it are less useful. We designed a TF-IDF-based weighting scheme to assign higher weights to more relevant CUIs and semantic types, and multiply these *W*_CUI_ to its corresponding **h**_*i*_:

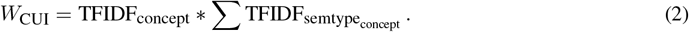

#### 4.1.3 Path Reasoning and Ranking

For each node representation **h**_*i*_, we used its n-hop 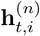 of the set neighborhood 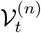 for **h**_*i*_ and the associated relation edge 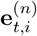 to generate the corresponding path embeddings:

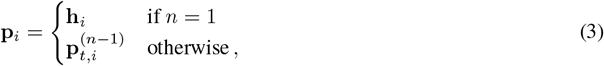

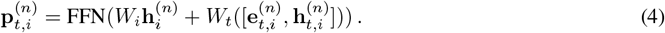

where FFN is feed-forward network, and *n* is the number of hop in the subgraph 𝒢_*src*_.

For each path embedding **p**_*i*_, we proposed two attention mechanisms, i.e., MultiHead attention (MultiAttn) and Trilinear attention (TriAttn), to compute its logical relation leveraging the input narrative representation **h**_x_ and input list of CUIs **h**_v_, both of which are encoded by SapBERT. We further defined **H**_*i*_ as context relevancy matrix, and **Z**_*i*_ as concept relevancy matrix:

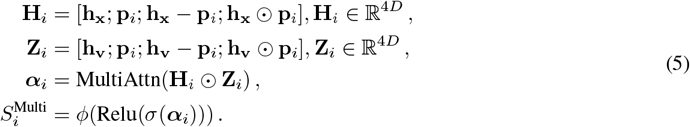

These relevancy matrices were inspired by prior work on natural language inference (Conneau et al., 2017), specifying the logical relations as matrix concatenation, difference, and product. An alternative design is Trilinear attention which learns the intricate relations by three attention maps:

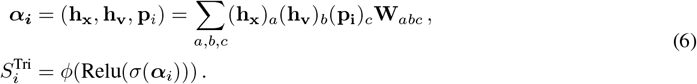

where **h**_x_, **p**_*i*_ and **h**_v_ have same dimensionality *D*, and *cp* is a MLP. Finally, we aggregated the MultiAttn or TriAttn scores on all candidate nodes, and select the top *N* entity 𝒱_*N*_ for next hop iteration based on the aggregated scores:

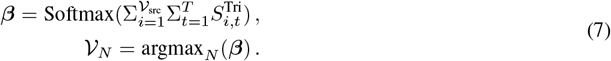

#### 4.1.4 Loss Function

Our loss function consisted of two parts, i.e., a CUI prediction loss and a contrastive learning loss:

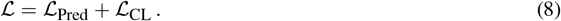

For prediction loss ℒ_Pred_, we used Binary Cross Entropy (BCE) loss to calculate whether selected 𝒱_*N*_ is in the gold label 𝒴:

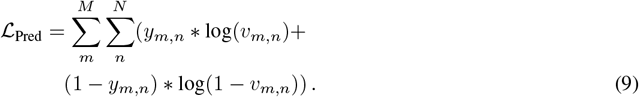

where *M* is the number of gold label 𝒴.

For contrastive learning loss ℒ_CL_, we encouraged the model to learn meaningful and discriminative representations by comparing with positive and negative samples:

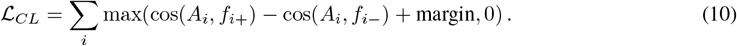

where *A*_*i*_ is the anchor embedding, defined as **h**_x_ ⨀ **h**_v_, and ⨀ is Hadamard product. Σ_*i*_ indicates a summation over a set of indices *i*, typically representing different training samples or pairs. Inspired from (Yasunaga et al., 2022), we construct cos(*A*_*i*_, *f*_*i*+_) and cos(*A*_*i*_, *f*_*i-*_) to calculate cosine similarity between *A*_*i*_ and positive feature *f*_*i*+_ or negative feature *f*_*i*_, respectively. This equation measures the loss when the similarity between an anchor and its positive feature is not significantly greater than the similarity between the same anchor and a negative feature, considering a margin for desired separation. Appendix C described the full Dr.Knows model training process.

#### 4.1.5 Prompting for foundational models

To incorporate graph model predicted paths into a prompt, we applied a prompt engineering strategy utilizing domain-independent prompt patterns, as delineated in White et al. (2023). Our prompt was constructed with two primary components: the *output customization* prompt, which specifies the requirement of exploiting knowledge paths, and the *context control patterns*, which are directly linked to the Dr.Knows’s output.

Given that our core objective was to assess the extent to which the prompt can bolster the model’s performance, it became imperative to test an array of prompts. Gonen et al. (2022) presented a technique, BetterPrompt, which relied on Selecting Prompts by Estimating Language Model Likelihood (spell). Essentially, we initiated the process with a set of manual task-specific prompts, subsequently expanding the prompt set via automatic paraphrasing facilitated by ChatGPT and backtranslation. We then ranked these prompts by their perplexity score (averaged over a representative sample of task inputs), ultimately selecting those prompts exhibiting the lowest perplexity.

Guided by this framework, we manually crafted five sets of prompts to integrate the path input, which are visually represented in Table 5. Specifically, the first three prompts were designed by a non-medical domain expert (computer scientist), whereas the final two sets of prompts were developed by a medical domain expert (a critical care physician and a medical informaticist). We designated the last two prompts as “Subject-Matter Prompts,” with the medical persona, and the first three prompts as “Non-Subject-Matter Prompts.” A comprehensive outline elucidating our approach to generating the prompt with paths can be found in Appendix E.

**Table 5:** Five manually designed prompts (Output Customization) and two path representation styles (Context Control) we create for the path-prompting experiments. There are 10 prompt patterns in total (5 Output Customization x 2 Context Control). For each Output Customization prompt, we generate 50 paraphrases using ChatGPT and run BETTERPROMPT to obtain the perplexity. This table also include the average perplexity for each prompt. Prompts with *are also deployed for path-less T5 fine-tuning (baseline).

| Group | Output Customization Prompts and No. | Perplexity |
| --- | --- | --- |
| Non-Subj. | A. <Explain> B. You may utilize these facts: C. You may find these facts helpful: | 3.86e-13 |
| Subj. | D. /E. *Act as a medical doctor, and list the top three direct and indirect diagnoses from the Assessment. *(You will be provided with some hints from a knowledge graph.) Explain the reasoning and assumptions behind your answer. | 1.03e-03 |
| No. | Context Control with Path Presentation |  |
| 1 | Structural.e.g. "*Infectious Diseases* -> *has pathological process* -> *Pneumonia*"<br>"*->*" is added to tokenizers as a new token. |  |
| 2 | Clause. e.g. "*Infectious Diseases has pathological process Pneumonia*" |  |

### 4.2 Experiments and Automated Evaluation

We trained the proposed Dr.Knows (TriAttn_*W*_ and MultiAttn_*W*_) on in-house and mimic dataset. We obtained a data split of 600, 81, and 87 on the mimic dataset and 3885, 520, 447 on the in-house dataset. The main task is to assess how well Dr.Knows predicts diagnoses using CUIs. To achieve this, we analyzed the text in the plan section using a concept extractor and extract the CUIs that fall under the semantic type T047 Disease and Syndromes. Specifically, we included the CUIs that are guaranteed to have at least one path with a maximum length of 2 hops between the target CUIs and input CUIs. These selected CUIs formed the “gold” CUI set, which was used for training and evaluating the model’s performance. Appendix B and D described the preprocessing and training setup, respectively.

Since Dr.Knows predicts the top *N* CUIs, we measured the Recall@N and Precision@N as below. The F-score is the harmonic mean between Recall and Precision, which will also be reported.

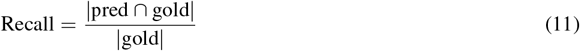

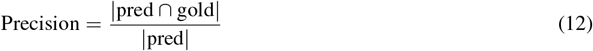

When evaluating the output diagnoses, we applied the above evaluation metric as well as ROUGE (Lin, 2004). Specifically, ROUGE is a widely used set of metrics designed for evaluating the quality of machine-generated text by comparing it to reference texts. We utilized the ROUGE-L variant, which is based on the longest common substring, and the ROUGE-2 variant, which focuses on bigram matching.

### 4.3 Metrics Development for Human Evaluation

#### 4.3.1 Motivation

Existing frameworks of human evaluation have been implemented for generative AI on certain tasks such as radiology report generation, but the field of diagnosis generation remains underdeveloped. Robust evaluation methodologies like SaferDX (Singh et al., 2019)have paved the way for assessing missed diagnostic opportunities, but their potential integration with Language Model evaluations has yet to be explored. Our refined framework underscores the pressing need for a structured human evaluation approach, which remains the reference standard and overcomes the limitations of quantitative evaluations. Our rigor in modeling SaferDx, performing a thorough literature review, and iterative user-centered design by subject matter experts helped to design an evaluation framework that was further validated by strong inter-rater agreement by medical experts.

We identified seven broad aspects widely deployed in human evaluation for biomedical NLP tasks: (1) Factual Consistency (Guo et al., 2020; Yadav et al., 2021; Wallace et al., 2020; Abacha et al., 2023; Moramarco et al., 2021; Otmakhova et al., 2022; Dalla Serra et al., 2022; Cai et al., 2022), (2) Hallucination (Guo et al., 2020; Umapathi et al., 2023), (3) Quality of Evidence (Otmakhova et al., 2022; Singhal et al., 2023), (4) Safety / Potential for Harm (Singhal et al., 2023; Dalla Serra et al., 2022; Adams et al., 2023), (5) Confidence (Otmakhova et al., 2022), (6) Omission (Abacha et al., 2023), and (7) Linguistic Quality (Radev and Tam, 2003; Guo et al., 2020). These aspects were then broken down and more clearly defined for inclusion in a human evaluation framework. The only factor not considered was Linguistic Quality. This factor was tied to general domain tasks and those intent on the fluency and readability of generated text for the general population. However, in a clinical setting, this is not a key focus so attention was given to aspects relating to content, instead.

#### 4.3.2 Survey Development

##### Evaluation criteria

The intent of evaluation of clinical diagnostic reasoning tasks is to verify that inclusion of generative LLMs in the clinical setting does not introduce additional potential for harm on patients. Therefore, the diagnostic evaluation portion was largely influenced by the revised SaferDx instrument (Singh et al., 2019) because of its applications in identifying and defining diagnostic errors and their potential for harm. Based on this instrument and our 6 identified aspects of manual evaluation from literature searching, the diagnostic evaluation process was broken down into 4 sections: (1) Accuracy, (2) plausibility, (3) specificity, and (4) omission and uncertainty. Accuracy was intended to capture the factuality of the diagnostic output as well as penalize a model for hallucinating output that does not qualify as a diagnosis. plausibility, which is conditional on Accuracy, was intended to capture the potential for harm present in an inaccurate diagnosis. specificity, which is conditional on plausibility, is defined as the level of detail provided in the diagnosis. Finally, omission and uncertainty defined cases when a diagnosis is not included in the list of outputted diagnoses but would be considered by a clinician in the clinical setting based upon the input data. In the case of the omission, the uncertainty further defined the reasons as *aleatoric uncertainty* – when LLM has been provided with the necessary information but has not utilized it; *epistemic uncertainty* – when the input to LLM does not contain the data needed to make a diagnosis.

The quality of evidence aspect of evaluation becomes a key factor in evaluating the reasoning output because clinical diagnostic reasoning is not a definitive process. Therefore, the reasoning evaluation portion was largely influenced by the framework established in (Singhal et al.2023), because of their rigorous validity measures compared to other established evaluation frameworks and focus on evidence quality as an aspect of evaluation. We utilized three of the aspects of their evaluation framework - (1) reading comprehension, (2) rationale, and (3) recall of knowledge - and incorporated an aspect on (4) omission of diagnostic reasoning. reading comprehension was intended to capture if a model understood the information in a progress note. rationale was intended to capture the inclusion of incorrect reasoning steps. recall of knowledge was intended to capture the hallucination of incorrect facts as well as the inclusion of irrelevant facts in the output. Finally, omission served the same purpose as previously by capturing when the model failed to support conclusions or provide evidence for a diagnostic choice.

In addition to the aspects outlined above, the evaluators were also asked to answer questions based on the amount of abstraction present in each part of the output. This was to ascertain how the knowledge paths influenced the type of output produced and whether or not the model was able to use abstraction. Since abstraction does not directly equate to better text generation, these questions did not impact the scoring process, but served as an additional piece of information. For the reasoning output, Effective Abstraction, conditional on abstraction, was also utilized to determine if any of the abstracted output aided or hindered the reasoning.

##### Implementation

Figure 11 presents the structure of the proposed human evaluation survey, and the questions asked under each scoring aspect. Each model output consists of the model predicted diagnoses (“*Diagnosis*”) and reasoning (“*<Reasoning>*“). We scored diagnoses and reasoning both at the individual instance level and their entirety. The scoring aspects of each component were highlighted in §4.3.2.

**Figure 11:**
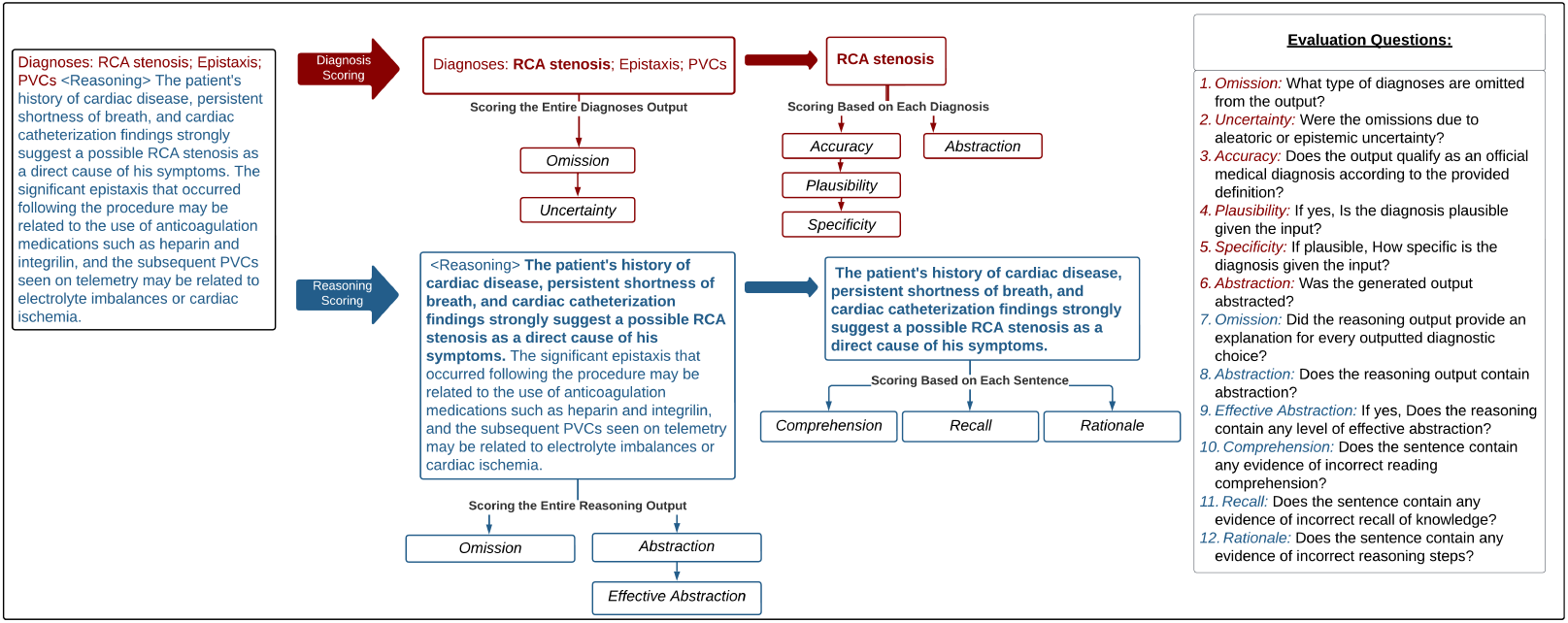
The structure of the survey questions given an LLM output. The output consists of two components: diagnoses (red-colored text) and reasoning (blue-colored text). For each component, there are corresponding questions evaluating certain aspects.

The evaluation framework was implemented utilizing the Research Electronic Data Capture (REDCap) web application. The input, output, and gold standards were auto-populated into REDCap for the evaluators. Each evaluator was treated as a different arm in a longitudinal data collection framework that had two defined events: one for the model utilizing knowledge graph paths and one for the model without them. The guidelines given to each evaluator contain a step-by-step guide on how to complete an evaluation in the REDCap system. We attached the complete survey and REDCap interface to Supplementary Materials.

##### Validation

We employ two crucial methods, construct validity and content validity, to ensure the robustness and effectiveness of our proposed human evaluation process. Construct validity and content validity are indispensable tools in the realm of research and assessment, playing pivotal roles in the verification of usability and the quality of our evaluation framework. two senior physicians who are experts with more than 10 years of experience in taking care of patients and also board-certified in clinical informatics served as advisors and pilot test users, which met the requirements for content validity. The helped design the user guide and train two medical professionals with medical school training to perform the human evaluations.

The construct validity is supported by the inter-annotator agreement between the two senior physicians and two medical professionals. Utilizing approximately 20 output examples from each model, iterative corrections were made to the human evaluation process to maximize usability, clarity, and applicability. Upon agreement between the clinicians, the two medical professionals were trained to complete the evaluations. They were trained on approximately 20 output examples from each model until they were in agreement with the senior clinicians (*Kappa >* 0.7). The inter-annotator agreement between the two final evaluators was also verified (*Kappa >* 0.7).

The construct validity of the proposed survey received further support from our literature search on previous work that used the same criteria or standards for assessment. We examined over 50 manual evaluation framework for text summarization from publications in the Association for Computational Linguistics and PubMed, and identified the 7 broad aspects of manual evaluation (see §**??**). We also used the SaferDx survey instrument to guide our survey development, ensuring the survey was designed with a focus on diagnostic safety.

#### 4.3.3 Survey scoring

Once the resident and medical student were verified as in agreement with the senior clinicians, each was given a set of output records from each model to evaluate. In total, at least 92 records were evaluated for each model.

##### Processing Steps

In the pre-processing phase, we handled missing values in the Plausibility and Specificity category differently depending on the cause.

Due to the inherent branching logic within some of the categories, missing values were substituted with a value of 0 during the score calculations. Additionally, we implemented a scoring transformation to the Comprehension, Recall, and Rationale questions: to address the reverse interpretation of these questions, we employed a transformation formula: (6 *- x*).

##### Diagnosis Scoring

The diagnosis score *D*_*i*_ given a record *i* is computed as below:

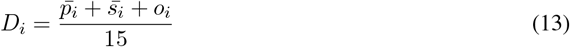

where 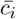 is the mean of the plausibility scores for record 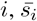 is the mean of the specificity scores for record *i, o*_*i*_ is the mean of the omission and uncertainty scores for record *i*, The denominator is 15 because each component was scored on a 5-point Likert scale and this 15 normalizes the scores into a (0, 1) scale.

##### Reasoning Scoring

The reasoning score *R*_*i*_ given a record *i* is computed as below:

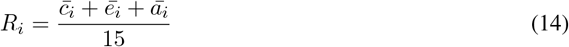

where 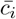 is the mean of the comprehension scores, 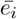 is the mean of the recall scores, 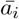 is the mean of the rationale scores for record *i*. The denominator is 15 because each component was scored on a 5-point Likert scale and this 15 normalizes the scores into a (0, 1) scale.

#### 4.3.4 Significance Testing

Statistical significance testing was performed utilizing a paired assumption. Since the KG and No KG scoring processes were done using the same progress notes, a pair was considered to be the score from each model for a particular progress note. Tests on statistical significance between normalized diagnosis and reasoning scores used a two-sided paired t-test. This is because the diagnosis and reasoning scores were quantitative values on a 0 to 1 scale. In cases where analysis was done on aspects of the scores (i.e. specificity, omission, plausibility), a McNemar test was utilized. The Likert and binary scale values were considered nominal categories for this test. All statistical significance testing was performed in R v4.3.1.

## Data Availability

All data produced in the present study are available upon reasonable request to the authors

## References

Asma Ben Abacha, Wen-wai Yim, George Michalopoulos, and Thomas Lin. 2023. An investigation of evaluation metrics for automated medical note generation. arXiv preprint 2305.17364.

Griffin Adams, Emily Alsentzer, Mert Ketenci, Jason Zucker, and Noémie Elhadad. 2021. What’s in a summary? laying the groundwork for advances in hospital-course summarization. In Proceedings of the conference. Association for Computational Linguistics. North American Chapter. Meeting, volume 2021, page 4794. NIH Public Access.

Griffin Adams, Jason Zucker, and Noémie Elhadad. 2023. A meta-evaluation of faithfulness metrics for long-form hospital-course summarization. arXiv preprint 2303.03948.

Claudio Aracena, Fabián Villena, Matías Rojas, and Jocelyn Dunstan. 2022. A knowledge-graph-based intrinsic test for benchmarking medical concept embeddings and pretrained language models. In Proceedings of the 13th International Workshop on Health Text Mining and Information Analysis (LOUHI), pages 197–206.

Akari Asai, Zeqiu Wu, Yizhong Wang, Avirup Sil, and Hannaneh Hajishirzi. 2023. Self-rag: Learning to retrieve, generate, and critique through self-reflection. arXiv preprint 2310.11511.

Erin P Balogh, Bryan T Miller, and John R Ball. 2015. Improving diagnosis in health care.

Christian Baumgartner. 2023. The potential impact of chatgpt in clinical and translational medicine. Clinical and translational medicine, 13(3).

Olivier Bodenreider. 2004. The unified medical language system (umls): integrating biomedical terminology. Nucleic acids research, 32(suppl_1):D267–D270.

PJ Brown, JL Marquard, B Amster, M Romoser, J Friderici, S Goff, and D Fisher. 2014. What do physicians read (and ignore) in electronic progress notes? Applied clinical informatics, 5(02):430–444.

Pengshan Cai, Fei Liu, Adarsha Bajracharya, Joe Sills, Alok Kapoor, Weisong Liu, Dan Berlowitz, David Levy, Richeek Pradhan, and Hong Yu. 2022. Generation of patient after-visit summaries to support physicians. In Proceedings of the 29th International Conference on Computational Linguistics, pages 6234–6247, Gyeongju, Republic of Korea. International Committee on Computational Linguistics.

Loredana Caruccio, Stefano Cirillo, Giuseppe Polese, Giandomenico Solimando, Shanmugam Sundaramurthy, and Genoveffa Tortora. 2024. Can chatgpt provide intelligent diagnoses? a comparative study between predictive models and chatgpt to define a new medical diagnostic bot. Expert Systems with Applications, 235:121186.

Alexis Conneau, Douwe Kiela, Holger Schwenk, Loïc Barrault, and Antoine Bordes. 2017. Supervised learning of universal sentence representations from natural language inference data. In Proceedings of the 2017 Conference on Empirical Methods in Natural Language Processing, pages 670–680, Copenhagen, Denmark. Association for Computational Linguistics.

Pat Croskerry. 2005. Diagnostic failure: A cognitive and affective approach. Advances in Patient Safety: From Research to Implementation (Volume 2: Concepts and Methodology).

Francesco Dalla Serra, William Clackett, Hamish MacKinnon, Chaoyang Wang, Fani Deligianni, Jeff Dalton, and Alison Q. O’Neil. 2022. Multimodal generation of radiology reports using knowledge-grounded extraction of entities and relations. In Proceedings of the 2nd Conference of the Asia-Pacific Chapter of the Association for Computational Linguistics and the 12th International Joint Conference on Natural Language Processing (Volume 1: Long Papers), pages 615–624, Online only. Association for Computational Linguistics.

Molla S Donaldson, Janet M Corrigan, Linda T Kohn, et al. 2000. To err is human: building a safer health system.

Luciano Floridi and Massimo Chiriatti. 2020. Gpt-3: Its nature, scope, limits, and consequences. Minds and Machines, 30:681–694.

Bryant Furlow. 2020. Information overload and unsustainable workloads in the era of electronic health records. The Lancet Respiratory Medicine, 8(3):243–244.

Yanjun Gao, Dmitriy Dligach, Timothy Miller, Matthew M Churpek, and Majid Afshar. 2023. Overview of the problem list summarization (probsum) 2023 shared task on summarizing patients’ active diagnoses and problems from electronic health record progress notes. arXiv preprint 2306.05270.

Yanjun Gao, Dmitriy Dligach, Timothy Miller, Dongfang Xu, Matthew MM Churpek, and Majid Afshar. 2022. Summarizing patients’ problems from hospital progress notes using pre-trained sequence-to-sequence models. In Proceedings of the 29th International Conference on Computational Linguistics, pages 2979–2991.

Hila Gonen, Srini Iyer, Terra Blevins, Noah A Smith, and Luke Zettlemoyer. 2022. Demystifying prompts in language models via perplexity estimation. arXiv preprint 2212.04037.

Yue Guo, Wei Qiu, Yizhong Wang, and Trevor Cohen. 2020. Automated lay language summarization of biomedical scientific reviews. CoRR, abs/2012.12573.

Bin He, Di Zhou, Jinghui Xiao, Xin Jiang, Qun Liu, Nicholas Jing Yuan, and Tong Xu. 2020. Bert-mk: Integrating graph contextualized knowledge into pre-trained language models. In Findings of the Association for Computational Linguistics: EMNLP 2020, pages 2281–2290.

Kung-Hsiang Huang, Mu Yang, and Nanyun Peng. 2020. Biomedical event extraction with hierarchical knowledge graphs. In Findings of the Association for Computational Linguistics: EMNLP 2020, pages 1277–1285, Online. Association for Computational Linguistics.

Alistair EW Johnson, Tom J Pollard, Lu Shen, Li-wei H Lehman, Mengling Feng, Mohammad Ghassemi, Benjamin Moody, Peter Szolovits, Leo Anthony Celi, and Roger G Mark. 2016. Mimic-iii, a freely accessible critical care database. Scientific data, 3(1):1–9.

Tomoyuki Kuroiwa, Aida Sarcon, Takuya Ibara, Eriku Yamada, Akiko Yamamoto, Kazuya Tsukamoto, and Koji Fujita. 2023. The potential of chatgpt as a self-diagnostic tool in common orthopedic diseases: Exploratory study. Journal of Medical Internet Research, 25:e47621.

Patrick Lewis, Ethan Perez, Aleksandra Piktus, Fabio Petroni, Vladimir Karpukhin, Naman Goyal, Heinrich Küttler, Mike Lewis, Wen-tau Yih, Tim Rocktäschel, et al. 2020. Retrieval-augmented generation for knowledge-intensive nlp tasks. Advances in Neural Information Processing Systems, 33:9459–9474.

Hao Li, Yuping Wu, Viktor Schlegel, Riza Batista-Navarro, Thanh-Tung Nguyen, Abhinav Ramesh Kashyap, Xiaojun Zeng, Daniel Beck, Stefan Winkler, and Goran Nenadic. 2023. Pulsar: Pre-training with extracted healthcare terms for summarising patients’ problems and data augmentation with black-box large language models. arXiv preprint 2306.02754.

Chin-Yew Lin. 2004. Rouge: A package for automatic evaluation of summaries. In Text Summarization Branches Out, pages 74–81.

Fangyu Liu, Ehsan Shareghi, Zaiqiao Meng, Marco Basaldella, and Nigel Collier. 2021. Self-alignment pretraining for biomedical entity representations. In Proceedings of the 2021 Conference of the North American Chapter of the Association for Computational Linguistics: Human Language Technologies, pages 4228–4238.

Jinghui Liu, Daniel Capurro, Anthony Nguyen, and Karin Verspoor. 2022. “note bloat” impacts deep learning-based nlp models for clinical prediction tasks. Journal of biomedical informatics, 133:104149.

Qiuhao Lu, Dejing Dou, and Thien Huu Nguyen. 2021. Parameter-efficient domain knowledge integration from multiple sources for biomedical pre-trained language models. In Findings of the Association for Computational Linguistics: EMNLP 2021, pages 3855–3865.

Potsawee Manakul, Yassir Fathullah, Adian Liusie, Vyas Raina, Vatsal Raina, and Mark Gales. 2023. Cued at probsum 2023: Hierarchical ensemble of summarization models. arXiv preprint 2306.05317.

Francesco Moramarco, Damir Juric, Aleksandar Savkov, and Ehud Reiter. 2021. Towards objectively evaluating the quality of generated medical summaries. arXiv preprint 2104.04412.

Sohn Nijor, Gavin Rallis, Nimit Lad, and Eric Gokcen. 2022. Patient safety issues from information overload in electronic medical records. Journal of Patient Safety, 18(6):e999–e1003.

Yulia Otmakhova, Karin Verspoor, Timothy Baldwin, and Jey Han Lau. 2022. The patient is more dead than alive: exploring the current state of the multi-document summarisation of the biomedical literature. In Proceedings of the 60th Annual Meeting of the Association for Computational Linguistics (Volume 1: Long Papers), pages 5098–5111, Dublin, Ireland. Association for Computational Linguistics.

Shirui Pan, Linhao Luo, Yufei Wang, Chen Chen, Jiapu Wang, and Xindong Wu. 2023. Unifying large language models and knowledge graphs: A roadmap. arXiv preprint 2306.08302.

Dragomir R. Radev and Daniel Tam. 2003. Summarization evaluation using relative utility. In Proceedings of the Twelfth International Conference on Information and Knowledge Management, CIKM ‘03, page 508–511, New York, NY, USA. Association for Computing Machinery.

Colin Raffel, Noam Shazeer, Adam Roberts, Katherine Lee, Sharan Narang, Michael Matena, Yanqi Zhou, Wei Li, and Peter J Liu. 2020. Exploring the limits of transfer learning with a unified text-to-text transformer. The Journal of Machine Learning Research, 21(1):5485–5551.

Maya Rotmensch, Yoni Halpern, Abdulhakim Tlimat, Steven Horng, and David Sontag. 2017. Learning a health knowledge graph from electronic medical records. Scientific reports, 7(1):5994.

Adam Rule, Steven Bedrick, Michael F Chiang, and Michelle R Hribar. 2021. Length and redundancy of outpatient progress notes across a decade at an academic medical center. JAMA Network Open, 4(7):e2115334–e2115334.

Guergana K Savova, James J Masanz, Philip V Ogren, Jiaping Zheng, Sunghwan Sohn, Karin C Kipper-Schuler, and Christopher G Chute. 2010. Mayo clinical text analysis and knowledge extraction system (ctakes): architecture, component evaluation and applications. Journal of the American Medical Informatics Association, 17(5):507–513.

Kurt Shuster, Spencer Poff, Moya Chen, Douwe Kiela, and Jason Weston. 2021. Retrieval augmentation reduces hallucination in conversation. In Findings of the Association for Computational Linguistics: EMNLP 2021, pages 3784–3803.

Hardeep Singh, Arushi Khanna, Christiane Spitzmueller, and Ashley ND Meyer. 2019. Recommendations for using the revised safer dx instrument to help measure and improve diagnostic safety. Diagnosis, 6(4):315–323.

Karan Singhal, Shekoofeh Azizi, Tao Tu, S Sara Mahdavi, Jason Wei, Hyung Won Chung, Nathan Scales, Ajay Tanwani, Heather Cole-Lewis, Stephen Pfohl, et al. 2023. Large language models encode clinical knowledge. Nature, 620(7972):172–180.

Luca Soldaini and Nazli Goharian. 2016. Quickumls: a fast, unsupervised approach for medical concept extraction. In MedIR workshop, sigir, pages 1–4.

Logesh Kumar Umapathi, Ankit Pal, and Malaikannan Sankarasubbu. 2023. Med-halt: Medical domain hallucination test for large language models. arXiv preprint 2307.15343.

Byron C. Wallace, Sayantan Saha, Frank Soboczenski, and Iain James Marshall. 2020. Generating (factual?) narrative summaries of rcts: Experiments with neural multi-document summarization. CoRR, abs/2008.11293.

Guojia Wan and Bo Du. 2021. Gaussianpath: A bayesian multi-hop reasoning framework for knowledge graph reasoning. In Proceedings of the AAAI conference on artificial intelligence, volume x35, pages 4393–4401.

Lawrence L Weed. 1969. Medical records, medical education, and patient care: The problem-oriented medical record as a basic tool. Cleveland, OH: Press of Case Western University.

Jules White, Quchen Fu, Sam Hays, Michael Sandborn, Carlos Olea, Henry Gilbert, Ashraf Elnashar, Jesse Spencer-Smith, and Douglas C Schmidt. 2023. A prompt pattern catalog to enhance prompt engineering with chatgpt. arXiv preprint 2302.11382.

Keyulu Xu, Weihua Hu, Jure Leskovec, and Stefanie Jegelka. 2019. How powerful are graph neural networks? In International Conference on Learning Representations.

Shweta Yadav, Deepak Gupta, Asma Ben Abacha, and Dina Demner-Fushman. 2021. Reinforcement learning for abstractive question summarization with question-aware semantic rewards. CoRR, abs/2107.00176.

Michihiro Yasunaga, Antoine Bosselut, Hongyu Ren, Xikun Zhang, Christopher D Manning, Percy S Liang, and Jure Leskovec. 2022. Deep bidirectional language-knowledge graph pretraining. Advances in Neural Information Processing Systems, 35:37309–37323.

Huaixiu Steven Zheng, Swaroop Mishra, Xinyun Chen, Heng-Tze Cheng, Ed H Chi, Quoc V Le, and Denny Zhou. 2023. Take a step back: Evoking reasoning via abstraction in large language models. arXiv preprint 2310.06117.

