## Supplemental File for "Large Language Models and Medical Knowledge Grounding for Diagnosis Prediction"

| |
|---|
| (Annotated Diagnoses and CUIs) |
| BRBPR; Pancreatitis; Cholecystitis |
| C0018932; C0008325; C0030305 |

Table 6: Annotated diagnoses and their corresponding Concept Unique Identifiers (CUIs)

#### A MIMIC-III and IN-HOUSE Data Examples

Table 1 describes the statistical information of MIMIC-III and IN-HOUSE datasets, where IN-HOUSE dataset includes all hospital settings compared to MIMIC-III dataset focusing on ICU. In addition, the average number of input and output CUIs on IN-HOUSE dataset is more than MIMIC-III ones, but MIMIC-III contains almost 50% abstractive concepts,i.e. the concepts or CUIs not being mentioned in the input, compared to IN-HOUSE with less than 1%. Table 7 shows plan section data examples from MIMIC-III and IN-HOUSE, where different diagnosis/problem is outlined and the corresponding treatment plan is given.

| |
|---|
| **MIMIC-III Plan section:**<br>1. GIB: s/p endoscopy Thursday without clear source of bleeding; Friday repeat EGD showed a duodenal ulcer with a clean base and no active bleeding. C. diff negative, h. pylori pending. Tagged red scan negative, CTA done and no read back.<br>- appreciate GI recommendations<br>- continue to trend Hct Q8hours, goal > 25, goal INR < 1.5 - f/up h. pylori<br>- f/up CTA read<br>2. FEVER: new fever yesterday with rigors, new productive cough and RLL infiltrate on CXR. Started on vanco/zosyn, CTA at lung bases looks like B atelectasis vs. PNA, not enough windows to evaluate fully. Sputum cx with oral flora.<br>- continue vanco + zosyn<br>- f/up cultures |
| **IN-HOUSE Plan section:**<br>NEURO: This patient is cognitively intact. Does not describe any pain.<br>REPIRATORY: As mentioned, the patient request to be do not intubate, have been giving him face mask to maintain saturation and will perform nonrebreather mask and BiPAP as needed.<br>CARDIOVASCULAR: As mentioned above, the patient this morning found to be profoundly hypotensive, with a SIRS picutre of tachypnea, elevated while bloob count, bandmia, and low tempretuare at .<br>GI: Recent diverticular and AVM bleed secondary to anticogulation. |

Table 7: MIMIC-III and IN-HOUSE plan section data examples.

#### B Datasets Preprocessing Steps

Here we describe how to extract ground truth CUIs for the CUI prediction task from a given input medical narrative and the target plan sections. We employ two medical concept extractors that rely on UMLS SNOMED CUIs: cTAKES, a Java toolkit (Savova et al., 2010), and QuickUMLS, a lightweight Python package (Soldaini and Goharian, 2016). For our IN-HOUSE dataset, we utilize cTAKES for note pre-processing, as it is integrated into a pre-built pipeline specifically designed for data analytics within the hospital's HIPAA-compliant network. On the other hand, for the MIMIC-III dataset, we opt for QuickUMLS due to its user-friendly nature, facilitating the reproducibility of the study on the public dataset. Both cTakes and QuickUMLS demonstrate similar levels of performance. Table 6 presents an example list of diagnoses and CUIs, after being parsed by cTAKES and QuickUMLS.

In order to create the ground truth for the CUI prediction task (§2.2), we aim to identify the set of CUIs within the plan sections that ensure the existence of the shortest path connecting them to the input CUIs. It is important to note that without such a path, a knowledge graph-based solution will be unable to locate the corresponding CUIs. Mathematically, the ground truth set of CUIs is defined as follows: For every CUI $c_i$ in the ground truth set, there exists a path with a maximum length of 2 to the CUI $c_j$, where $c_j$ is an element of the input set of CUIs. We exclude paths with lengths exceeding 2 (i.e., more than 2 hops) from consideration, as our empirical findings suggest that source and target concepts connected by 3-hop paths have a lower likelihood of being clinically meaningful.

In practice, for every pair of CUIs $(c_i, c_j)$, where $c_i$ is in the plan sections CUIs and $c_j$ in the input text, we run a Dijkstra algorithm to find if there exists a path with length as 2 or less. Algorithm 1 presents the workflow of identifying the ground truth CUIs for CUI prediction tasks.

**Algorithm 1** Extract Path-based CUIs as Ground Truth for CUI Prediction Task

**procedure** FINDSHORTESTPATHS(PlanSectionCUIs, InputCUIs)
  Initialize TargetOutput as an empty set
  **for each** concept $ci$ **in** PlanSectionCUIs **do**
    **for each** concept $cj$ **in** InputCUIs **do**
      Run Dijkstra's algorithm to find the shortest paths from $ci$ to $cj$
      Let $shortestPath$ be the shortest path found, $l$ be the length of the $shortestPath$
      **if** $l \leq 2$ **then**
        Include $ci$ in TargetOutput
      **else**
        Ignore $ci$
      **end if**
    **end for**
  **end for**
  **return** TargetOutput
**end procedure**

#### C Details of DR.KNOWS Graph Model Training Algorithm

**Algorithm 2** DR.KNOWS Graph Model Training for Path Selection

**Require:** UMLS knowledge graph $\mathcal{G}$, input patient text $\mathbf{x}$
Build $\mathcal{V}_{\text{src}}$ and $\mathcal{E}_{\text{src}}$ given $\mathbf{x}$
Initialise CUI embedding CUI($\mathbf{v}_i$) or CUI$(\mathbf{v}_i)_{\text{W}}$ as $\mathbf{h}_i$ and relation embedding $\mathbf{e}_t$ as one-hot embedding
**while** not halting **do**
  **if** $t > 1$ **then** Update $\mathcal{V}_{\text{src}}$
  **end if**
  Update node representation of $\mathbf{v}_i$ based on Eq. 1
  Generate path embedding $\mathbf{p}_i$ in the Path Encoder based on Eq. 3-4
  Rank path embedding using MultiAttn or TriAttn based on Eq. 5-7
**end while**
Calculate loss function based on Eq. 8-10
Update model's parameter $\boldsymbol{\theta}$

The DR.KNOWS model whole training for path selection is shown in Algo. 2

#### D DR.KNOWS Training Setup

For the proposed graph models, we use SapBERT (Liu et al., 2021) as an encoder to encode input patient text and CUI concepts. Within the graph model components in Figure 10, we use a Transformer as the path encoder, and MultiAttn or TriAttn is used as the path ranker. The number of $t$ is 2. At each hop iteration during training, the selected top $N$ is 8. Then $N$ is set as 4 or 6 for inference. The oracle setting is True in the graph model training, and then set as False for inference. The AdamW optimizer with 1e-5 learning rate is used by decaying it every 3 epochs during training. The graph model is trained for 10 epochs.

### E Paraphrased Customization Prompts using ChatGPT

We provide manual task-specific prompts in Table 5 to ChatGPT, and ask it to paraphrase those prompts by giving an instruction as “Paraphrase this prompt <Manual Prompts> to 50 different prompts”.[1] Then BETTERPROMPT (Gonen et al., 2022) is used to evaluate those prompts providing perplexity, and top 5 prompts with the lowest perplexity are selected for further finetuning. Table 8 shows the full list of top 5 "Non-Subject-Matter" and "Subject-Matter Prompts" paraphrased prompts with the corresponding perplexity scores.

**ChatGPT Zero-Shot Prompting Setup** We employ the same prompt selection approach as previously stated, that is, to pick the prompt with the lowest perplexity. ChatGPT is sensitive to the prompt. Following the originally selected prompt, ChatGPT does not separate its diagnosis and reasoning generation, as the original prompt does not include specific output requirements. Consequently, we initially evaluate the prompt using 73 samples from the PROBSUM development set. Subsequently, we proceed with the zero-shot experiments on the test set, while making additional adjustments to the prompt to enable ChatGPT to generate diagnoses and reasoning separately. Specifically, we ask ChatGPT to use “<Reasoning>” to separate its diagnoses and assumption. We utilize the same development set to fine-tune the hyperparameters essential for text generation. To ensure consistency and reproducibility, we set the temperature to 1 and impose a maximum limit of 160 tokens for the generated output. This limit prevents excessive generation and the introduction of extraneous information or noise.

Another key observation during the setup of the zero-shot prompt-based experiment is that ChatGPT solely copies the output from knowledge paths without considering the input note. This is because the prompt does not explicitly require ChatGPT to utilize information from the input note. This deviation from our original intention to test if path prompts improve the note summarization task of foundation models undermines the primary focus on the note as the main source of information. Therefore, we add additional instructions to prompt ChatGPT to focus on the input note and be aware of the noisy knowledge graphs.

[1] https://openai.com/blog/chatgpt

| Manual Prompts | Top 5 Paraphrased Variants | Perplexity |
|---|---|---|
| You may utilize these facts: | Employ these facts to bolster your understanding: | 2.95e-8 |
| | Employ these facts to strengthen your research: | 3.02e-8 |
| | Employ these facts to broaden your knowledge: | 3.47e-8 |
| | Use these facts to complement your findings: | 3.59e-8 |
| | Use these facts to broaden your perspective: | 5.10e-8 |
| You may find these facts helpful: | Consider these facts as potentially practical: | 2.26e-9 |
| | Consider these facts as potentially insightful: | 3.50e-9 |
| | Consider these facts as potentially beneficial: | 4.66e-9 |
| | Consider these facts as potentially valuable: | 4.68e-9 |
| | You might find these facts advantageous: | 4.72e-9 |
| Act as a medical doctor, and list the top three direct and indirect diagnoses from the Assessment. Explain the reasoning and assumptions behind your answer. | Imagine yourself as a physician and outline the three key direct and indirect diagnoses identified in the Assessment. Discuss the reasoning and assumptions that guide your response. | 3.98e-4 |
| | In the role of a physician, generate a list of the Assessment's three primary direct and indirect diagnoses. Shed light on the reasoning and assumptions that inform your decision. | 5.50e-4 |
| | Act as a medical doctor and detail the top three direct and indirect diagnoses resulting from the Assessment. Justify your choices by explaining the reasoning and assumptions involved. | 5.81e-4 |
| | In your role as a medical doctor, please outline the primary three direct and indirect diagnoses based on the Assessment. Elucidate the reasoning and assumptions underlying your choices. | 5.83e-4 |
| | Take on the role of a doctor and indicate the Assessment's top three direct and indirect diagnoses. Justify your choices by explaining the underlying reasoning and assumptions. | 6.08e-4 |
| Act as a medical doctor, and list the top three direct and indirect diagnoses from the input note. You will be provided with some hints from a knowledge graph. Explain the reasoning and assumptions behind your answer. | Imagine you are a medical professional equipped with a knowledge graph, and generate the top three direct and indirect diagnoses from the input note. | 3.47e-4 |
| | Imagine you are a physician incorporating knowledge graph hints, and outline the top three direct and indirect diagnoses stemming from the input note. Discuss the reasoning and assumptions that inform your diagnostic analysis. | 4.69e-4 |
| | Act as a doctor, leveraging insights from a knowledge graph, and compile a list of the primary direct and indirect diagnoses derived from the input note. Explain the reasoning and assumptions underlying your responses. | 1.04e-3 |
| | In the capacity of a physician, employ the information from a knowledge graph to identify the three principal direct and indirect diagnoses stemming from the input note. Justify your answers by explaining the reasoning and assumptions involved. | 1.10e-3 |
| | Imagine yourself as a doctor utilizing a knowledge graph, and provide a comprehensive breakdown of the top three direct and indirect diagnoses resulting from the input note. Clarify the reasoning and assumptions that underpin your diagnostic selections. | 1.17e-3 |

Table 8: The full list of top 5 "Non-Subject-Matter" and "Subject-Matter Prompts" paraphrased prompts with the corresponding perplexity scores (Note: as most <Explain> prompt variants generated by ChatGPT are irrelevant to path prompting compared to the other manual prompt variants. We only evaluate the <Explain> itself using BETTERPROMPT, and its perplexity is 3.86e-13)
.

#### Manual Evaluation Guidelines and Workflow

##### Introduction

Generative AI has made monumental progress in recent years. Their utilization in the clinical setting has the potential to revolutionize the clinical decision-making process. The core elements of clinical diagnostic reasoning are the ability to gather, understand and integrate clinical evidence, reason over the evidence using medical knowledge, and summarize relevant diagnoses. These cognitive skills are mapped to the following cNLP research areas: (1) medical knowledge representation, (2) clinical evidence understanding and integration, and (3) diagnosis generation and summarization [7]. Both knowledge representation and clinical experience are used simultaneously in an interactive fashion by clinicians and serve as the design for artificial intelligence systems to model. Thus far, evaluation of these systems has not undergone consistent rigorous evaluation presenting a lack of thoroughly tested and verified success in a clinical setting. This manual evaluation intends to cover the important aspects of the diagnostic process in a way that increases inter-annotator agreement and becomes a building block for the development of evaluation in the area.

For this project, a generative AI model is prompted to imagine it is a medical professional in order to determine the diagnoses for a patient given an input note. The system is also provided with examples for how to approach the problem before being given the input. An example of the prompts can be found in Appendix 21. The input notes for this project come from MIMIC-III. It incorporates the assessment section and subjective section from real patient daily progress notes across multiple intensive care units. The assessment section presents important information about the patient, their reason for hospitalization, and any other relevant information. This is followed by the subjective section after the tag <Subjective> which includes the Chief Complaint, 24 Hour Events, and Allergies of the patient. There will be two two different versions of the input: one that only includes the items mentioned above and another that includes potentially relevant knowledge paths [6]. The other part of the input, the knowledge paths, are generated based upon the MIMIC III information and utilizes the UMLS semantic network to identify the important concepts and their relevant relations with other medical concepts. These paths start with a concept which is connected to another by a joining phrase (e.g., Procedure (procedure) --> temporally follows --> Graft Versus Host Diseases). These graphs are then read hierarchically from right to left (e.g., Graft Versus Host Disease temporally follows Procedure). These connections are generated by a separate model outside this project and therefore can contain incorrect information, bad reasoning, or other mistakes. Therefore, an evaluator should rely more on their own medical background for evaluation and only utilize the knowledge graphs when accurate or helpful.

Note: Make sure to review this entire document before beginning the evaluation process

**Definitions and Evaluation Goals**

- According to the National Library of Medicine Unified Medical Language System [5] has defined ***diagnosis*** as the following:
    - (1) MESH definition: The determination of the nature of a disease or condition, or the distinguishing of one disease or condition from another. Assessment may be made through physical examination, laboratory tests, or the likes. Computerized programs may be used to enhance the decision-making process.
    - (2) NCI: The investigation, analysis and recognition of the presence and nature of disease, condition, or injury from expressed signs and symptoms; also, the scientific determination of any kind; the concise results of such an investigation. A diagnosis is made from the study of the signs and symptoms of a disease.
    - UMLS Semantic Types: Disease or Syndrome
    - In your evaluation of Diagnosis Yes/No - use the UMLS Metathesaurus Browser to help with determining if the term is a diagnosis by using guidance for concept unique identifiers (CUIs) that have semenative type for 'disease or syndrome.'

- ***Differential Diagnosis*** is defined as the determination of which one of two or more diseases or conditions a patient is suffering from by systematically comparing and contrasting results of diagnostic measures. Among the accepted concepts in the UMLS metathesaurus, a differential can include an established diagnosis, admitting diagnosis, principal diagnosis, working diagnosis, secondary diagnosis, prior diagnosis, suspected diagnosis, early diagnosis, uncertain diagnosis, postmortem diagnosis, referral diagnosis, transfer diagnosis, gross diagnosis, ED diagnosis, improbable diagnosis, missed diagnosis, delayed diagnosis, late diagnosis, etc. The goal of this evaluation provides an evaluation for plausible principle/primary and secondary diagnoses. Also, to evaluate missed and improbable diagnoses. [5]

- A diagnosis is ***plausible*** if it is not contradicted by any information in the input and would be included as a potential diagnosis in the differential diagnosis process
    - Note: When answering questions based upon plausibility, "Strongly Disagree" indicates that a diagnosis is so implausible that it has the potential to cause harm, create bias, or negatively impact the patient's care. If the diagnosis is likely just incorrect but not potentially harmful then just mark "Disagree".
    - Example: It is not plausible to assign prostate cancer as a potential diagnosis when the patient was assigned female at birth

- A diagnosis is ***specific*** if the level of detail provided in the diagnosis. A diagnosis can be too broad, where the diagnosis ignores information from the input that would imply a diagnosis that is more granular or abstract a more granular version of the problem, or very narrow where the diagnosis is as granular as possible.
    - Examples:

| Very General | General | Neutral | Specific | Highly specific |
|---|---|---|---|---|
| Lung Disease | Acute respiratory failure | Pneumonia | Viral Pneumonia | COVID Pneumonia with ARDS |
| Fever of Unknown origin | Infection | MRSA Infection | MRSA Bacteremia | Line-associated MRSA Bacteremia |

- A diagnosis is considered ***omitted*** if is not included in the list of outputted diagnoses, but would be considered by a clinician in the clinical setting based upon the input data to the LLM.
    - Example:

| | |
|---|---|
| Input | [System Prompt and Few-Shot Examples, See Appendix]<br>61 year old woman with newly [**Hospital 5068**] [**Hospital **] transfered from BMT unit on<br>day 18 s/p induction (7+3) chemotherapy, with febrile neutropenia and tachypnea. TITLE: Chief Complaint: 61 year old woman with newly diagosed AML transfered from BMT unit on day 18 s/p induction (7+3) chemotherapy, with febrile neutropenia and tachypnea. 24 Hour Events: MULTI LUMEN - START 08:15 PM from the floor BLOOD CULTURED - At 09:00 PM BLOOD CULTURED - At 04:53 AM FEVER - 104.0 F - 08:15 PM Allergies: Penicillins Rash; Sulfa (Sulfonamide Antibiotics) Rash; Hydrochlorothiazide Rash; |
| Output | Diagnoses: Febrile neutropenia; Chemotherapy-induced pneumonia; Sepsis |
| Gold Standard | Respiratory distress; Fever: Possible etiologies include neutropenia, pulmonary infection (? viral infection on CT GGO), typhlitis, diverticulitis (both seen on<br>CT abdomen), drug fever, leukemia; Thrombocytopenia: Secondary to recent induction chemo; Anemia: Likely secondary to leukemia and recent chemotherapy; |

- A diagnosis is ***direct*** if it
  - is the primary diagnosis/problem listed for hospitalization and available in the input to the LLM
  - is a problem/diagnosis related to the primary signs/symptoms in the input to the LLM

- A diagnosis is ***indirect*** if it
  - is a complication/subsequent event or organ failure related to the primary diagnosis/problem
  - is another listed diagnosis/problem from the overall progress note that is not part of the primary diagnosis/problem
  - is a diagnosis/problem that is not previously mentioned but closely related (i.e., same organ system) to the primary diagnoses/problems

- Example – Direct Diagnosis & Indirect Diagnosis

| | |
|---|---|
| Input | [System Prompt and Few-Shot Examples, See Appendix]<br>61 year old woman with newly [**Hospital 5068**] [**Hospital **] transfered from BMT unit on<br>day 18 s/p induction (7+3) chemotherapy, with febrile neutropenia and tachypnea. TITLE: Chief Complaint: 61 year old woman with newly diagosed AML transfered from BMT unit on day 18 s/p induction (7+3) chemotherapy, with febrile neutropenia and tachypnea. 24 Hour Events: MULTI LUMEN - START 08:15 PM from the floor BLOOD CULTURED - At 09:00 PM BLOOD CULTURED - At 04:53 AM FEVER - 104.0 F - 08:15 PM Allergies: Penicillins Rash; Sulfa (Sulfonamide Antibiotics) Rash; Hydrochlorothiazide Rash; |
| Gold Standard | Respiratory distress; Fever: Possible etiologies include neutropenia, pulmonary infection (? viral infection on CT GGO), typhlitis, diverticulitis (both seen on<br>CT abdomen), drug fever, leukemia; Thrombocytopenia: Secondary to recent induction chemo; Anemia: Likely secondary to leukemia and recent chemotherapy; |

- In the case of this project an omission due to ***aleatoric uncertainty*** results when the model has been provided with the necessary information, but has not utilized it. The human evaluator can deduce the diagnosis but the model was not able to (i.e., inherent limitation of the model and not the input data). IF the Gold Standard contains a diagnosis that is also apparent from the input data THEN this is aleatoric.

- In the case of this project an omission due to ***epistemic uncertainty*** results when the input to the model does not contain the data needed to make a diagnosis. The human evaluator would also not be able to deduce a diagnosis without more information (i.e., inherent limitation of the data input itself). IF the Gold Standard contains a diagnosis that is NOT apparent from the input data THEN this is epistemic.

  - Note: The uncertainty type can be determined by comparing the omissions, gold standards, and input to determine if the model has been given the opportunity to make the correct diagnosis

- Generated Text is considered ***abstracted*** when the output creates new phrases and sentences that relay the most useful information from the original text [1]. For this project, a diagnosis is only considered an abstraction if it does not appear in the input data, but does in the output diagnoses. So *extractive* summarization is if the input data mentions a disease like 'COVID pneumonia' and the output diagnosis also provided 'COVID pneumonia'. *Abstractive* summarization is if the input data describes renal failure with bacteremia and the output diagnosis states 'Sepsis' (in this case the model infers sepsis correctly from a set of findings).

- Generated Text is considered ***extracted*** when it involves pulling key phrases from the source document and combining them to make an output without any additional changes or inclusions [1]. See example above of extractive summarization.

    - Example – Abstracted Text & Extracted Text

| | |
|---|---|
| Input | [System Prompt and Few-Shot Examples, See Appendix]<br>61 year old woman with newly [**Hospital 5068**] [**Hospital **] transfered from BMT unit on<br>day 18 s/p induction (7+3) chemotherapy, with febrile neutropenia and tachypnea. TITLE: Chief Complaint: 61 year old woman with newly diagosed AML transfered from BMT unit on day 18 s/p induction (7+3) chemotherapy, with febrile neutropenia and tachypnea. 24 Hour Events: MULTI LUMEN - START 08:15 PM from the floor BLOOD CULTURED - At 09:00 PM BLOOD CULTURED - At 04:53 AM FEVER - 104.0 F - 08:15 PM Allergies: Penicillins Rash; Sulfa (Sulfonamide Antibiotics) Rash; Hydrochlorothiazide Rash; |
| Output | Diagnoses: Febrile neutropenia; Chemotherapy-induced pneumonia; Sepsis<br><br>< Reasoning> The patient has febrile neutropenia and tachypnea, which could suggest the presence of an infection. The multiple blood cultures drawn also support this possibility. Chemotherapy-induced pneumonia is a common complication in patients undergoing chemotherapy for leukemia. Additionally, with a fever of 104.0 F, sepsis should be considered as a differential diagnosis. |

- ***Reading comprehension*** refers to the ability of the model to understand the input it has been given.
    - Example of Incorrect Reading Comprehension:
    - The model reasons that Epistaxis is a direct diagnosis because the patient had significant epistaxis following NGT placement, however the model has failed to utilize the fact that epistaxis was noted as resolved in the subjective section of the note or that the cause of the epistaxis has already been noted.

**General Procedures**

1. Begin the Manual Evaluation Section with Step I: Diagnosis Scoring
   - ***Example Output***:
     Diagnoses: Febrile neutropenia; Chemotherapy-induced pneumonia; Sepsis
     < Reasoning> The patient has febrile neutropenia and tachypnea, which could suggest the presence of an infection. The multiple blood cultures drawn also support this possibility. Chemotherapy-induced pneumonia is a common complication in patients undergoing chemotherapy for leukemia. Additionally, with a fever of 104.0 F, sepsis should be considered as a differential diagnosis.
2. Step Ia will be completed for each individual diagnosis listed in the output
   - ***Example Cont.*** Diagnoses: Febrile neutropenia; Chemotherapy-induced pneumonia; Sepsis
3. Only continue to the next question when an arrow indicates to do so based on your answer to the current question - Note: not every question will be answered for every diagnosis
   - ***Example Cont.***
     Diagnosis: Febrile neutropenia

| Question | Yes | No |
|---|---|---|
| Does the output qualify as an official medical diagnosis according to the provided definition? | ■ | |

| Question | 1: Strongly Disagree | 2: Disagree | 3: Neutral | 4: Agree | 5: Strongly Agree |
|---|---|---|---|---|---|
| Is the diagnosis plausible given the input? | | | | | ■ |

| Question | 1: Very General | 2: General | 3: Neutral | 4: Specific | 5: Very Specific |
|---|---|---|---|---|---|
| How specific is the diagnosis given the input? | | | ■ | | |

| Question | Yes | No |
|---|---|---|
| Was the generated output abstracted? | | ■ |

Diagnosis: Chemotherapy-induced pneumonia

| Question | Yes | No |
|---|---|---|
| Does the output qualify as an official medical diagnosis according to the provided definition? | ■ | |

| Question | 1: Strongly Disagree | 2: Disagree | 3: Neutral | 4: Agree | 5: Strongly Agree |
|---|---|---|---|---|---|
| Is the diagnosis plausible given the input? | | | | | ■ |

| Question | 1: Very General | 2: General | 3: Neutral | 4: Speci fic | 5: Very Specific |
|---|---|---|---|---|---|
| How specific is the diagnosis given the input? | | | | | ■ |

| Question | Yes | No |
|---|---|---|
| Was the generated output abstracted? | ■ | |

Diagnosis: Sepsis

| Question | Yes | No |
|---|---|---|
| Does the output qualify as an official medical diagnosis according to the provided definition? | ■ | |

| Question | 1: Strongly Disagree | 2: Disagree | 3: Neutral | 4: Agree | 5: Strongly Agree |
|---|---|---|---|---|---|
| Is the diagnosis plausible given the input? | | | | | ■ |

| Question | 1: Very General | 2: General | 3: Neutral | 4: Speci fic | 5: Very Specific |
|---|---|---|---|---|---|
| How specific is the diagnosis given the input? | | | | ■ | |

| Question | Yes | No |
|---|---|---|
| Was the generated output abstracted? | ■ | |

4. Repeat step Ia until all diagnoses in the output have been scored before moving on to the omission scoring
5. Complete step Ib based on the entire list of outputted diagnoses
   - Note: If multiple types of diagnoses have been omitted select the option that reflects the worst omission
   - ***Example Cont.***

| Question | 1: All Direct | 2: Majority Direct | 3: Majority Indirect | 4: All Indirect | 5: No Omissions |
|---|---|---|---|---|---|
| What type of diagnoses are omitted from the output? | | | | | c |

| Question | 1: All Epistemic | 2: Majority Epistemic | 3: Majority Aleatoric | 4: All Aleatoric | 5: No Omissions |
|---|---|---|---|---|---|
| Were the omissions due to aleatoric or epistemic uncertainty? | | | | | |

6. Continue on to Step II: Reasoning Scoring
   - ***Example Cont.*** < Reasoning> The patient has febrile neutropenia and tachypnea, which could suggest the presence of an infection. The multiple blood cultures drawn also support this possibility. Chemotherapy-induced pneumonia is a common complication in patients undergoing chemotherapy for leukemia. Additionally, with a fever of 104.0 F, sepsis should be considered as a differential diagnosis.

7. Step IIa will be completed for each individual sentence in the reasoning output
    - ***Example Cont.*** The patient has febrile neutropenia and tachypnea, which could suggest the presence of an infection.

| Question | 1: Strongly Disagree | 2: Disagree | 3: Neutral | 4: Agree | 5: Strongly Agree |
|---|---|---|---|---|---|
| Does the sentence contain any evidence of incorrect reading comprehension? (Indicating the input has not been understood) [4] | | | | | |
| Does the sentence contain any evidence of incorrect recall of knowledge? (Mention of an irrelevant and/or incorrect fact for answering the question) [4] | | | | | |
| Does the sentence contain any evidence of incorrect reasoning steps? (Incorrect rationale for a diagnostic choice) [4] | | | | | |

8. Answer all the three questions for each sentence
    - ***Example Cont.*** The multiple blood cultures drawn also support this possibility.

| Question | 1: Strongly Disagree | 2: Disagree | 3: Neutral | 4: Agree | 5: Strongly Agree |
|---|---|---|---|---|---|
| Does the sentence contain any evidence of incorrect reading comprehension? (Indicating the input has not been understood) [4] | | | | | |
| Does the sentence contain any evidence of incorrect recall of knowledge? (Mention of an irrelevant and/or incorrect fact for answering the question) [4] | | | | | |
| Does the sentence contain any evidence of incorrect reasoning steps? (Incorrect rationale for a diagnostic choice) [4] | | | | | |

- ***Example Cont.*** Chemotherapy-induced pneumonia is a common complication in patients undergoing chemotherapy for leukemia.

| Question | 1: Strongly Disagree | 2: Disagree | 3: Neutral | 4: Agree | 5: Strongly Agree |
|---|---|---|---|---|---|
| Does the sentence contain any evidence of incorrect reading comprehension? (Indicating the input has not been understood) [4] | | | | | |
| Does the sentence contain any evidence of incorrect recall of knowledge? (Mention of an irrelevant and/or incorrect fact for answering the question) [4] | | | | | |
| Does the sentence contain any evidence of incorrect reasoning steps? (Incorrect rationale for a diagnostic choice) [4] | | | | | |

9. Repeat step IIa until all sentences in the output have been scored
   - ***Example Cont.***Additionally, with a fever of 104.0 F, sepsis should be considered as a differential diagnosis.

| Question | 1: Strongly Disagree | 2: Disagree | 3: Neutral | 4: Agree | 5: Strongly Agree |
|---|---|---|---|---|---|
| Does the sentence contain any evidence of incorrect reading comprehension? (Indicating the input has not been understood) [4] | | | | | |
| Does the sentence contain any evidence of incorrect recall of knowledge? (Mention of an irrelevant and/or incorrect fact for answering the question) [4] | | | | | |
| Does the sentence contain any evidence of incorrect reasoning steps? (Incorrect rationale for a diagnostic choice) [4] | | | | | |

10. Complete step IIb based on the entire reasoning output
    - ***Example Cont.***

| Question | Yes | No |
| --- | --- | --- |
| Did the reasoning output provide an explanation for every generated diagnosis? | | |

| Question | Yes | No |
| --- | --- | --- |
| Does the reasoning output contain abstraction? | | |

| Question | Yes | No |
| --- | --- | --- |
| Does the reasoning contain any level of effective abstraction? | | |

**RedCap Specific Procedures**

Note: The records for each input/output combo up for evaluation have been imported into RedCap as records. Thus, an evaluator only needs to edit the created records rather than create new ones.

1. Utilizing your RedCap access point, navigate to the "Generative AI Qualitative Evaluation" project
2. Upon opening the project, click the "Add / Edit Records" tab on the left menu bar
3. The following page will be the entry point for every manual evaluation (there are 228*2 for this project)
4. Begin an evaluation by selecting a record that you have not yet completed
    a. There will be multiple evaluators for this project. Select the arm corresponding to you before selecting a record

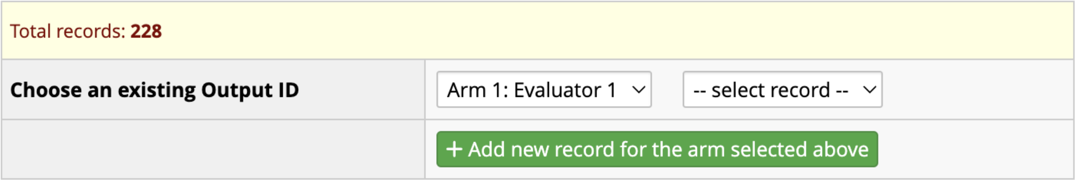


5. The record homepage will appear as shown below

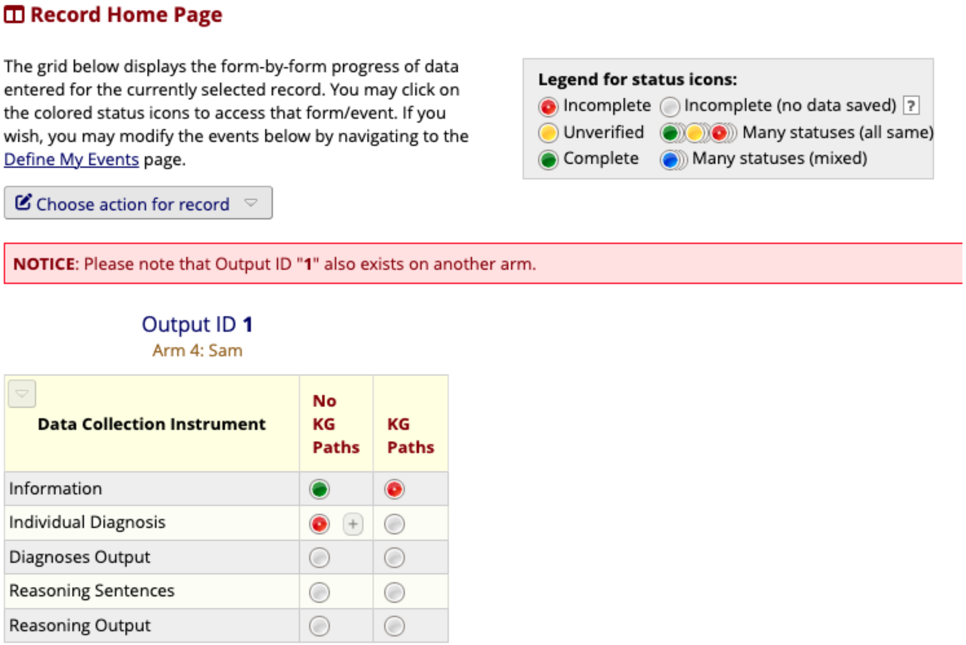


6. Each Output ID can have multiple events which correspond to the large language models being evaluated. They will be very similar, but contain slight differences based upon the individual model.
7. Evaluation must be completed twice for each Output ID – once per event

8. Each evaluation consists of 5 instruments representing the different aspects of the evaluation, you will need to complete all of them to complete an evaluation
9. Start with the Information instrument by clicking the red status circle under the ChatGPT event
10. Answer the questions for this instrument
    a. Note: The Input, Gold Standard, and Output will auto populate on every page for the evaluator's reference
11. Once you have completed an instrument change the status to complete before continuing to the next one

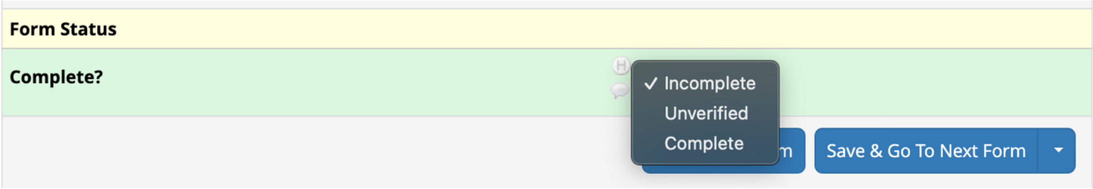


12. Continue to the next instrument by pressing the "Save & Go To Next Form" button at the bottom of the page
13. The "Individual Diagnosis" and "Reasoning Sentences" instruments will need to be repeated for each instance of a diagnosis/sentence in the provided output. These instruments will have the following information displayed at the top of the screen to inform you how many times to repeat the instrument
    a. Note: Each "Instance" is one completion of the instrument

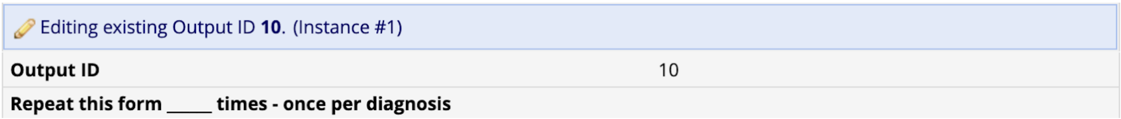


14. To repeat the instrument, press the blue down arrow and select "Save & Go To Next Instance"

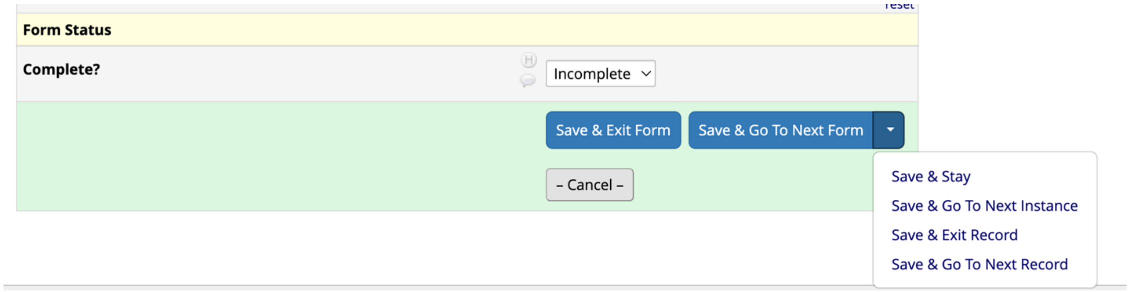


15. Once the instrument has been repeated the correct number of times, select "Save & Go To Next Form" to continue to the next instrument
16. Once you reach the final instrument, select "Save & Exit Form" which will return you to the record homepage to start the evaluation for the Llama2 event by repeating steps 9 - 16 for the Llama2 event
17. Once both events have been completed, return to the "Add / Edit Records" tab and select the next record to complete
18. If at any time you need to leave the evaluation and return later, press the "Save & Exit Form" button. Upon returning, you can select the same record and pick up where you left off

19. To check if you completed an evaluation make sure that all the status symbols for the record are green. You will be able to see the status for each instrument and each instance on the status page for that Output ID record

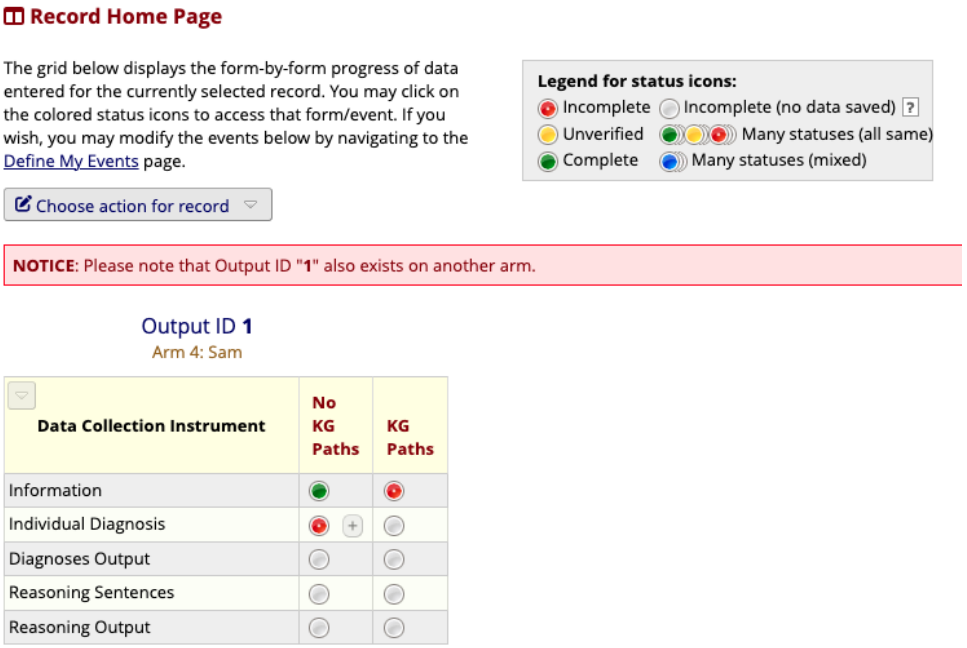


20. You are also able to view all the records that you have/need to complete through the "Record Status Dashboard" tab and navigating to the arm that represents you. This page can also be used to switch between different records by clicking the Output ID number to see the homepage for that record or by selecting any of the status circles to see the particular instrument/instance/event of a record

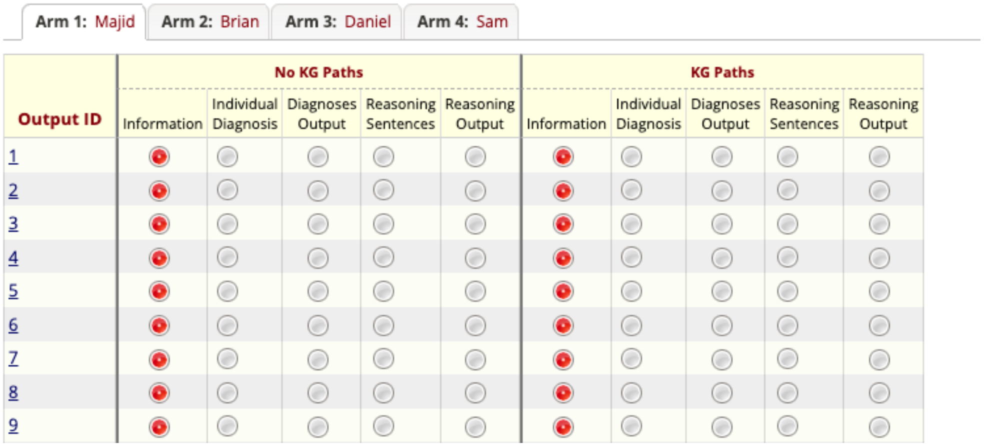

**Manual Evaluation Framework**

I. Diagnosis Scoring

a. Scoring Per Diagnosis Listed in Output

| Question | Yes | No |
|---|---|---|
| Does the output qualify as an official medical diagnosis according to the provided definition? | | |

| Question | 1: Strongly Disagree | 2: Disagree | 3: Neutral | 4: Agree | 5: Strongly Agree |
|---|---|---|---|---|---|
| Is the diagnosis plausible given the input? | | | | | |

| Question | 1: Very General | 2: General | 3: Neutral | 4: Speci fic | 5: Very Specific |
|---|---|---|---|---|---|
| How specific is the diagnosis given the input? | | | | | |

| Question | Yes | No |
|---|---|---|
| Was the generated output abstracted? | | |

b. Omission Scoring Based on Entire List of Diagnoses in Output

| Question | 1: All Direct | 2: Majority Direct | 3: Majority Indirect | 4: All Indirect | 5: No Omissions |
|---|---|---|---|---|---|
| What type of diagnoses are omitted from the output? | | | | | |

| Question | 1: All Epistemic | 2: Majority Epistemic | 3: Majority Aleatoric | 4: All Aleatoric | 5: No Omissions |
|---|---|---|---|---|---|
| Were the omissions due to aleatoric or epistemic uncertainty? | | | | | |

II. Reasoning Scoring

a. Scoring Per Sentence in Reasoning Output

| Question | 1: Strongly Disagree | 2: Disagree | 3: Neutral | 4: Agree | 5: Strongly Agree |
|---|---|---|---|---|---|
| Does the sentence contain any evidence of incorrect reading comprehension? (Indicating the input has not been understood) [4] | | | | | |
| Does the sentence contain any evidence of incorrect recall of knowledge? (Mention of an irrelevant and/or incorrect fact for answering the question) [4] | | | | | |
| Does the sentence contain any evidence of incorrect reasoning steps? (Incorrect rationale for a diagnostic choice) [4] | | | | | |

b. Omission Scoring for Whole Reasoning Output

| Question | Yes | No |
|---|---|---|
| Did the reasoning output provide an explanation for every outputted diagnostic choice? | | |

| Question | Yes | No |
|---|---|---|
| Does the reasoning output contain abstraction? | | |

| Question | Yes | No |
|---|---|---|
| Does the reasoning contain any level of effective abstraction? | | |

#### References


[1] Ecosystem (LEDU) E. Medium. 2018 [cited 2023 Sep 5]. A Quick Introduction to Text Summarization in Machine Learning. Available from: https://towardsdatascience.com/a-quick-introduction-to-text-summarization-in-machine-learning-3d27ccf18a9f

[2] Hüllermeier, E., Waegeman, W. Aleatoric and epistemic uncertainty in machine learning: an introduction to concepts and methods. *Mach Learn* **110**, 457–506 (2021). https://doi.org/10.1007/s10994-021-05946-3

[3] National Academies of Sciences, Engineering, and Medicine. 2015. Improving Diagnosis in Health Care. Washington, DC: The National Academies Press. https://doi.org/10.17226/21794.

[4] Singhal K, Azizi S, Tu T, Mahdavi SS, Wei J, Chung HW, et al. Large language models encode clinical knowledge. Nature. 2023 Jul 12;1–9.

[5] UMLS Metathesaurus Browser [Internet]. [cited 2023 Sep 5]. Available from: https://uts.nlm.nih.gov/uts/umls; The definitions for Diagnosis and Differential Diagnosis are pulled from the UMLS Metathesaurus and Semantic Network because the knowledge paths that were generated for the input on this project were based off of the same knowledge base

[6] Gao Y, Li R, Caskey J, Dligach D, Miller T, Churpek MM, et al. Leveraging A Medical Knowledge Graph into Large Language Models for Diagnosis Prediction [Internet]. arXiv; 2023 [cited 2023 Sep 8]. Available from: http://arxiv.org/abs/2308.14321

[7] Gao Y, Dligach D, Miller T, Caskey J, Sharma B, Churpek MM, et al. DR.BENCH: Diagnostic Reasoning Benchmark for Clinical Natural Language Processing. Journal of Biomedical Informatics. 2023 Feb;138:104286.

**Appendix A: No Path Prompt Template (contains persona/system, instruction, and 5-shot examples)**

Imagine you are a medical professional, and generate the top three direct and indirect, differential diagnoses from the input note.

Example 1: A 60 year old woman with recurrent ALL with CNS involvement s/p Omaya removal due to VRE contamination & SDH evacuation. She is now doing well and awake s/p extubation, afebrile and her WBC count is trending downward. <Subjective> NGT placed/TF started
Femoral line changed over wire by IR
c diff x 2 negative, 3rd pending
d/c'ed gentamicin per ID
Sulfa (Sulfonamides)
Rash;
Penicillins
Rash;
Latex
Hives; Wheezing
Red Dye
"   Headache; ""thra"
Darvon (Oral) (Propoxyphene Hcl)
Nausea/Vomiting
Percodan (Oral) (Oxycodone Hcl/Oxycodon Ter/Asa)
Nausea/Vomiting
Aspirin
Nausea/Vomiting
Aspartame
Unknown;
Fentanyl
Arrhythmia/Palp
f
Review of systems is unchanged from admission except as noted below
Review of systems:

Use # to separate the output diagnoses, then write a separate text that starts with '<Reasoning>' to explain the reasoning behind your answer:

Diagnoses: CNS VRE; LEUKOCYTOSIS; ALL

Example 2: 79yo F with dCHF (75%, [**3-26**]), CAD (RCA stent '[**04**], LCx BMS in '[**23**]),
a-fib, HTN, DM2, ESRD on HD, cryptogenic cirrhosis with variceal bleeding (s/p banding '[**25**]) and LGIB (divericulosis, angiectasia '[**23**]) admitted with intertrochanteric femoral fracture after fall at home s/p ORIF c/b post-op hypotension and is transfered to MICU for close

observation. <Subjective> BP in 70s while asleep 500 cc IVF with return to baseline in 90s
Keflex (Oral) (Cephalexin Monohydrate)
severe rash;
Heparin Agents
Thrombocytopeni
f
Review of systems is unchanged from admission except as noted below
Review of systems:

Use # to separate the output diagnoses, then write a separate text that starts with '<Reasoning>' to explain the reasoning behind your answer:

Diagnoses: Intermittent, post-op hypotension; Infiltrate on CXR; Thrombocytopenia:; Cyrptogenic cirrhosis c/b variceal bleed:; DM2

Example 3: 53 yoM w/ a h/o schizoaffective disorder presents s/p fall with atrial flutter with a rapid ventricular response, intracranial lesion, and lung mass who has new dx of squamous cell lung cancer with extension into left atrium, and was started on IV amio load overnight. <Subjective> sinus tachy to 140s overnight ( 5:30am), gave IV verapamil
- Onc-neurosurg / Dr. [**First Name8 (NamePattern2) 164**] [**Last Name (NamePattern1) 2506**] to see in morning
- dry on exam, negative balance so given 1L IVF overnight
- completed Levo/Flag Abx ( 10d) today
- got in touch with [**Doctor Last Name **], he is coming in tomorrow & can address goals of care more
- now switched over to PO amiodarone
- Atrovent nebs added
- foley placed back in, incontinence & need to monitor UOP
No Known Drug Allergies
Review of systems is unchanged from admission except as noted below
Review of systems:

Use # to separate the output diagnoses, then write a separate text that starts with '<Reasoning>' to explain the reasoning behind your answer:

Diagnoses: Atrial & Ventricular Ectopy; L hilar mass/Brain Mass; Hypoxic respiratory failure; Weakness over right UE/LE; Schizoaffective disorder

Example 4: H/O CVA (STROKE, CEREBRAL INFARCTION), HEMORRHAGIC
.H/O RESPIRATORY FAILURE, CHRONIC
.H/O VENTRICULAR TACHYCARDIA, SUSTAINED
58 M with COPD s/p trach, h/o Nocardia, growing Aspergillus fumigatus on sputum cx, s/p recent thalamic CVA presents with hypoxic respiratory failure and found to have right middle lobe artery pulmonary embolus
. <Subjective> - Patient able to wean off vent to trach collar in AM of [**3-22**]

- Restarted on home dose of methadone
- Tube feeds restarted
- Started on treatment dose bactrim for stenotrophomonas/xanthomonas
- Spoke to LTAC physician to give update, has case management concerns with insurance, per case manager, will need to rescreen for LTAC
- CT head showing expected evolution of right thalamic hematoma, no new neuro recs

History obtained from Patient
History obtained from PatientNo Known Drug Allergies
f
Review of systems is unchanged from admission except as noted below
Review of systems:

Use # to separate the output diagnoses, then write a separate text that starts with '<Reasoning>' to explain the reasoning behind your answer:

Diagnoses: Hypoxic respiratory failure; Recent right thalamic hemorrhagic CVA

Example 5: 45 year old man with pmh significant for type I DM, ESRD on hemodialysis, labile blood pressure, presenting with hypertensive emergency. <Subjective> Started on labetolol drip. BP dropped from SBP 200s to 170s in first 20 minutes. Patient's mentation improved. BP at 2220- 144/61.
Resting comfortably- stable. Paged overnight for SBP continuing to be in the 170s. Gave patient 10mg of hydralazine at 1AM. BP stable there overnight. Paged at [**Pager number 10061**] on [**10-6**] saying BP in low 90s and patient dizzy. Gave 250cc NS bolus.
No Known Drug Allergies
Changes to and
f
Review of systems is unchanged from admission except as noted below
Review of systems:

Use # to separate the output diagnoses, then write a separate text that starts with '<Reasoning>' to explain the reasoning behind your answer:

Diagnoses: # Hypertensive emergency; Chest Pain; ESRD on HD; DM

**Appendix B: Graph Path Prompt Template (contains persona/system, instruction, and 5-shot examples, knowledge paths sourced from graph neural network)**

Imagine you are a medical professional equipped with a knowledge graph, and generate the top three direct and indirect, differential diagnoses from the input note. Try to extract direct diagnoses from input note, and prioritize the diagnoses directly inferred from the presentation of the problems in input note first. Keep in mind that the knowledge graph may include noisy or irrelevant information, only utilize the knowledge paths when you cannot find diagnoses from input notes and think most relevant and necessary.

Example 1: A 60 year old woman with recurrent ALL with CNS involvement s/p Omaya removal due to VRE contamination & SDH evacuation.  She is now doing well and awake s/p extubation, afebrile and her WBC count is trending downward. <Subjective> NGT placed/TF started
Femoral line changed over wire by IR
c diff x 2 negative, 3rd pending
d/c'ed gentamicin per ID
Sulfa (Sulfonamides)
Rash;
Penicillins
Rash;
Latex
Hives; Wheezing
Red Dye
"   Headache; ""thra"
Darvon (Oral) (Propoxyphene Hcl)
Nausea/Vomiting
Percodan (Oral) (Oxycodone Hcl/Oxycodon Ter/Asa)
Nausea/Vomiting
Aspirin
Nausea/Vomiting
Aspartame
Unknown;
Fentanyl
Arrhythmia/Palp
f
Review of systems is unchanged from admission except as noted below
Review of systems:
These are the knowlede paths: Arrhythmia --> self --> Arrhythmia --> self-->Arrhythmia <path> Redness --> self --> Redness --> self-->Redness <path> White blood cell count --> interprets --> Leukocytosis --> self-->Leukocytosis <path> Structure of femoral artery --> has finding site --> sinus tachycardia --> self-->sinus tachycardia <path> Oral cavity structure (body structure) --> has finding site --> PAROTITIS --> self-->PAROTITIS <path> Arrhythmia --> isa --> Cardiac arrest --> self-->Cardiac arrest

Use # to separate the output diagnoses, then write a separate text that starts with '<Reasoning>' to explain the reasoning behind your answer:

Diagnoses: CNS VRE; LEUKOCYTOSIS; ALL

Example 2: 79yo F with dCHF (75%, [**3-26**]), CAD (RCA stent '[**04**], LCx BMS in '[**23**]),
a-fib, HTN, DM2, ESRD on HD, cryptogenic cirrhosis with variceal
bleeding (s/p banding '[**25**]) and LGIB (divericulosis, angiectasia '[**23**])
admitted with intertrochanteric femoral fracture after fall at home s/p
ORIF c/b post-op hypotension and is transfered to MICU for close
observation. <Subjective> BP in 70s while asleep 500 cc IVF with return to baseline in 90s
Keflex (Oral) (Cephalexin Monohydrate)
severe rash;
Heparin Agents
Thrombocytopeni
f
Review of systems is unchanged from admission except as noted below
Review of systems:
These are the knowlede paths: systemic arterial hypertension --> self --> systemic arterial hypertension --> self-->systemic arterial hypertension <path> Unspecified chronic renal failure --> possibly equivalent to --> Renal failure: [chronic] or [end stage] --> possibly equivalent to-->Unspecified chronic renal failure <path> Thrombocytopenia --> has definitional manifestation --> thrombocytopenia --> self-->thrombocytopenia <path> Oral cavity structure (body structure) --> has finding site --> PAROTITIS --> self-->PAROTITIS <path> Unspecified chronic renal failure --> possibly equivalent to --> Renal failure: [chronic] or [end stage] <path> Thrombocytopenia --> has definitional manifestation --> thrombocytopenia --> isa-->Pancytopenia

Use # to separate the output diagnoses, then write a separate text that starts with '<Reasoning>' to explain the reasoning behind your answer:

Diagnoses: Intermittent, post-op hypotension; Infiltrate on CXR; Thrombocytopenia:; Cyrptogenic cirrhosis c/b variceal bleed:; DM2

Example 3: 53 yoM w/ a h/o schizoaffective disorder presents s/p fall with atrial
flutter with a rapid ventricular response, intracranial lesion, and
lung mass who has new dx of squamous cell lung cancer with extension
into left atrium, and was started on IV amio load overnight. <Subjective> sinus tachy to 140s overnight ( 5:30am), gave IV verapamil
- Onc-neurosurg / Dr. [**First Name8 (NamePattern2) 164**] [**Last Name (NamePattern1) 2506**] to see in morning
- dry on exam, negative balance so given 1L IVF overnight
- completed Levo/Flag Abx ( 10d) today
- got in touch with [**Doctor Last Name **], he is coming in tomorrow & can address goals of care more

- now switched over to PO amiodarone
- Atrovent nebs added
- foley placed back in, incontinence & need to monitor UOP
No Known Drug Allergies
Review of systems is unchanged from admission except as noted below
Review of systems:
These are the knowlede paths: Cardiac atrium --> has finding site --> auricular fibrillation --> self-->auricular fibrillation <path> Cardiac atrium --> has finding site --> AF - Paroxysmal atrial fibrill --> self-->AF - Paroxysmal atrial fibrill <path> Nasal sinus --> has finding site --> Obstructive sinusitis --> self-->Obstructive sinusitis <path> Cardiac atrium --> has finding site --> Atrial standstill --> self-->Atrial standstill <path> Incontinence --> self --> Incontinence --> self-->Incontinence <path> Cardiac atrium --> has finding site --> Chronic atrial fibrillation --> self-->Chronic atrial fibrillation

Use # to separate the output diagnoses, then write a separate text that starts with '<Reasoning>' to explain the reasoning behind your answer:

Diagnoses: Atrial & Ventricular Ectopy; L hilar mass/Brain Mass; Hypoxic respiratory failure; Weakness over right UE/LE; Schizoaffective disorder

Example 4: H/O CVA (STROKE, CEREBRAL INFARCTION), HEMORRHAGIC
.H/O RESPIRATORY FAILURE, CHRONIC
.H/O VENTRICULAR TACHYCARDIA, SUSTAINED
58 M with COPD s/p trach, h/o Nocardia, growing Aspergillus fumigatus
on sputum cx, s/p recent thalamic CVA presents with hypoxic respiratory
failure and found to have right middle lobe artery pulmonary embolus
. <Subjective> - Patient able to wean off vent to trach collar in AM of [**3-22**]
- Restarted on home dose of methadone
- Tube feeds restarted
- Started on treatment dose bactrim for stenotrophomonas/xanthomonas
- Spoke to LTAC physician to give update, has case management concerns
with insurance, per case manager, will need to rescreen for LTAC
- CT head showing expected evolution of right thalamic hematoma, no new
neuro recs
History obtained from Patient
History obtained from PatientNo Known Drug Allergies
f
Review of systems is unchanged from admission except as noted below
Review of systems:
These are the knowlede paths: Chronic (qualifier value) --> has course --> HYPERACTIVE AIRWAY DISEASE --> self-->HYPERACTIVE AIRWAY DISEASE <path> [D]Respiratory failure (situation) --> self --> [D]Respiratory failure (situation) --> self-->[D]Respiratory failure (situation) <path> STROKE --> self --> STROKE --> self-->STROKE <path> ventricular tachycardia (V-tach) --> self --> ventricular tachycardia (V-tach) --> self-->ventricular tachycardia (V-tach) <path> VENTRICULAR TACHYCARDIA --> self --> VENTRICULAR TACHYCARDIA --> self-->VENTRICULAR TACHYCARDIA <path> Chronic (qualifier

value) --> has course --> HYPERACTIVE AIRWAY DISEASE --> isa-->PULMONARY EMPHYSEMAS

Use # to separate the output diagnoses, then write a separate text that starts with '<Reasoning>' to explain the reasoning behind your answer:

Diagnoses: Hypoxic respiratory failure; Recent right thalamic hemorrhagic CVA

Example 5: 45 year old man with pmh significant for type I DM, ESRD on hemodialysis, labile blood pressure, presenting with hypertensive emergency. <Subjective> Started on labetolol drip. BP dropped from SBP 200s to 170s in first 20 minutes. Patient's mentation improved. BP at 2220- 144/61.
Resting comfortably- stable. Paged overnight for SBP continuing to be
in the 170s. Gave patient 10mg of hydralazine at 1AM. BP stable there
overnight. Paged at [**Pager number 10061**] on [**10-6**] saying BP in low 90s and patient dizzy. Gave 250cc NS bolus.
No Known Drug Allergies
Changes to and
f
Review of systems is unchanged from admission except as noted below
Review of systems:
These are the knowlede paths: Unspecified chronic renal failure --> self --> Unspecified chronic renal failure --> self-->Unspecified chronic renal failure <path> Pager --> self --> Pager --> self-->Pager <path> Unspecified chronic renal failure --> isa --> Chronic progressive renal failure --> self-->Chronic progressive renal failure <path> Admission to hospital --> self --> Admission to hospital --> self-->Admission to hospital <path> yr --> self --> yr --> self-->yr <path> NUM --> self --> NUM --> self-->NUM

Use # to separate the output diagnoses, then write a separate text that starts with '<Reasoning>' to explain the reasoning behind your answer:

Diagnoses: # Hypertensive emergency; Chest Pain; ESRD on HD; DM
